# SARS-CoV-2 Aerosol Transmission Indoors: A Closer Look at Viral Load, Infectivity, the Effectiveness of Preventive Measures and a Simple Approach for Practical Recommendations

**DOI:** 10.1101/2021.11.04.21265910

**Authors:** Martin Kriegel, Anne Hartmann, Udo Buchholz, Janna Seifried, Sigrid Baumgarte, Petra Gastmeier

**Author notes:** **Address for correspondence:** Prof. Dr.-Ing. Martin Kriegel, Hermann-Rietschel-Institut, TU Berlin. <u>Author contributions</u>: MK analyzed the data, developed the simplifications and performed the calculation. UB, SB supplied detailed data for retrospective analysis. PG, UB, JS and SB evaluated the results from different medical point of views. AH performed the literature research and the review of the models and calculations. MK and AH drafted the manuscript. All authors contributed to the interpretation of the results, critically revised the paper and agreed on the final version for submission.

## Abstract

Currently, airborne transmission is seen as the most important transmission path for SARS-CoV-2. In this investigation, a classic dose-response model is used on the one hand to find out retrospectively the probable viral load of the infectious source patient at the time of transmission in 25 documented outbreaks. We showed that an infection due to airborne transmission at a distance from the infectious person was probably only possible in the 25 outbreaks examined, with attack rates of 4-100%, if the viral load had been higher than 1E+08 viral copies/ml. This demonstrates that the viral load estimated from the swab might overestimate a person’s infectivity via aerosol, because a person is generally considered infectious, independent of the transmission way, when the viral load from the swab is 1E+06 viral copies/ml.

On the other hand, a possible approach is presented to predict the probable situational Attack Rate (PAR_s_) of a group of persons in a room through aerosol particles emitted by an infectious source patient. Four main categories of influence on the risk of infection are formed: First the emitted viruses, depending on the viral load and the amount of respiratory particles, and necessary number of reproducible viruses for infection, second the room-specific data and duration of stay of the group of people, third the activity of the exposed persons, and fourth the effect of personal protection (e.g. wearing masks from infectious and/or susceptible person).

Furthermore, a simplified method is presented to calculate either the maximum possible number of persons in a room, so that probably a maximum of one person becomes infected when an infectious person is in the room, or the PAR_s,simple_ for a given number of persons, ventilation rate and time of occupancy. We additionally show, taking into account organizational preventive measures, which person-related virus-free supply air flow rates are necessary to keep the number of newly infected persons to less than 1. The simple approach makes it easy to derive preventive organizational and ventilation measures. Our results show that the volume flow rate or a person-related flow rate is a much more effective parameter to evaluate ventilation for infection prevention than the air change rate. We suggest to monitor the CO_2_ concentration as an easy to implement and valid measurement system for indoor spaces.

Finally, we show that of the three measures, besides of wearing masks and increasing ventilation, testing contributes the most to the joint protective effect. This corresponds to the classic approach to implement protection concepts: preventing the source from entering the room and emitting viruses at all. In summary, a layered approach of different measures is recommended to mutually compensate for possible failures of any one measure (e.g. incorrect execution of tests, incorrect fit of masks or irregular window opening), to increase the degree of protection and thus reduce the risk of transmission of SARS-CoV-2.

## Introduction

The respiratory route is the main mode of transmission for the virus causing COVID-19 (SARS-CoV-2) [1, 2, 3, 4]. The virus is transported on particles that can enter the respiratory tract. Whereas larger particles (droplets) are only able to stay in the air for a short time and just in the near field (short range; approx. 1.5 m), because they settle down quickly, smaller particles (called aerosol particles; few μm until approximately 50 μm) are also concentrated in the near field but can also follow the air flow and cause therefore infections in the near and far field. Epidemiologically, short-range transmission (through droplets or aerosol particles) is distinguished from long-range transmission (aerosol particles) [5].

In order to perform an infection risk assessment for the airborne transmission in the far field and to introduce appropriate preventive measures, it would be necessary to know the amount of aerosol particles produced by an infected person during various activities, how many viruses stick to the particles and how many viruses are necessary to cause an infection.

However, this information is usually available very late during the course of a pandemic, if it can be determined at all. Another well-known approach is to use retrospective analysis of infection outbreaks that are very probably due to aerosol transmission of a single source patient to determine the unknown parameters inhaled virus copies and necessary copies to cause an infection.

### State of the Art

The so-called aerosol particles (liquid or solid particles suspended in a gas) and droplets differ by size. Particles, aerosol particles as well as macroscopic droplets, can be removed from the air by two different mechanisms: (i) air change because of a mechanical or natural ventilation and (ii) deposition on surfaces. In case of the investigation of amplifiable microorganisms transported on particles inactivation can also be seen as a removal method from air, because after the inactivation the microorganisms are not harmful anymore.

The air change rate can be calculated regarding equation (1) as the ratio of the volume flow (Q) to the room volume (V).

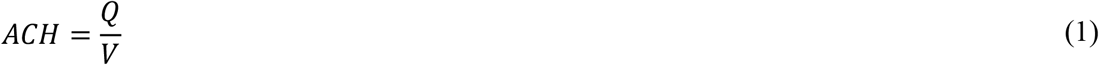

Typical air change rates for indoor environments range between 0.5 1/h for residential buildings and 8 1/h for occupied rooms with several attendants like meeting rooms in offices [6]. Often an air change rate of 6 air changes per hour is recommended to minimize the risk of infection [7]. Therefore, the air change rates advised for hospitals are used [6, 8].

Nevertheless, these recommendations are not based on infection prevention, but on CO_2_-emission of room occupants as well as general air quality requirements or thermal loads within the room.

The deposition rate for particles depends on their size, the air velocity, the turbulence intensity of the air movement and the ratio of surface area to room volume [9, 10, 11]. Thatcher et al [10] investigated the deposition of particles on surfaces depending on the room furniture and the air speed. The deposition rate increased for larger particles, higher air speed or an increased surface area of furniture. Particles between 0.55 and 8.66 μm were considered in this study.

Similar results were also found by Offerman [11], but for somewhat smaller particles (between 0.09 and 1.25 μm). Still the deposition rate increased for larger particles. For particles between 0.3 and 5 μm, which is considered to be the most important size range for airborne particles in equilibrium state, which will stay in air for a long time and are able to carry viruses, the deposition rate ranges between 0.1 and 0.4 1/h.

Different authors published results from measurements of the viral load in swabs of infected persons [12, 13, 14, 15]. The results show that on average a viral load of approximately 1E+06 viral copies per ml can be measured in the days before symptom onset for the wild-type and the Alpha variant. In some patients a viral load of up to 1E+12 viral copies per ml was also found around symptom onset. The temporal dynamic of the viral load depends on the course of the infection. Shortly before or at the onset of symptoms infected persons carry the highest viral load (peak load). For approximately 10% of the infected persons a raised peak load of ≥ 1E+08 viral copies per ml was found. Within approximately 24 hours, the viral load can increase by a factor of about 100. Whether a given patient is infectious can be estimated through measuring the probability to culture the virus. In [13] it is described that a viral load of 1E+06, 1E+07 and 1E+08 viral copies per ml taken from the swab have a culture probability in a lab of 20%, 50% and 75%, respectively. With the delta variant, the situation is somewhat different. The mean viral load is around 1E+08 viral copies per ml, significantly higher viral loads were found and the viral load decreases significantly more slowly after the peak [16, 17, 18, 19].

Particles in exhaled air are generated in the respiratory tract. The mucus is aerosolized so that the viruses contained in the mucus are distributed to the aerosol particles formed. The higher the viral load, the more particles actually carry virus. The aerosol particles disperse in the room air. People in the room inhale the aerosol particles. A direct correlation between culture viability in the laboratory via a swab and culture viability via aerosol particles cannot be drawn, so that it is also not possible to conclude that a person is infectious above a certain viral load.

The inactivation time depends on the pathogen and has therefore to be considered separately for each virus or bacteria investigated. For SARS-CoV-2 the inactivation time has been investigated experimentally by van Doremalen et al [20] as well as Dabisch et al [21]. Van Doremalen et al. investigated the inactivation in air as well as on different surfaces and compared it to values for SARS-CoV-1. The measured half-life of SARS-CoV-2 in air of approximately 1.1-1.2 h leads to an inactivation rate of approximately 0.6 1/h. Dabisch et al measured the decay rate of SARS-CoV-2 in air under different environmental conditions (temperature, humidity as well as simulated sunlight). The influence of simulated sunlight and temperature was found to be much larger than the influence of humidity, still all aspects have significant influence on the decay rate.

In 1978, Riley et al. [22] evaluated a measles outbreak in a suburban elementary school. Based on the number of susceptible persons (S), which have been infected (D) during each stage of infection of the source patient, the risk (P) for an infection at this stage has been calculated using equation (2). The risk for an infection has been defined as the percentage of infected persons from the number of pupils not already infected or vaccinated.

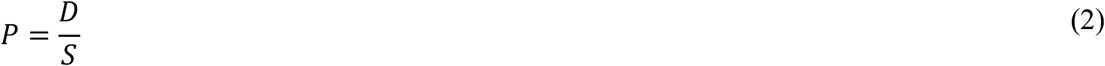

A Poisson-distribution of the risk of infection has been assumed as well as a stationary and evenly distributed concentration of the pathogens in the room air. Equation (3) shows the Poisson-distribution.

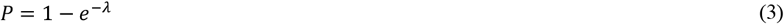

In 1955 Wells [23] defined a size called quantum as the number of emitted infectious units, where the probability to get infected is 1 − *e*^-1^ = 63.2%. Hence, a quantum can be seen as a combination of the amount of emitted virus-laden aerosol particles and a critical dose, which may result in an infection in 63.2 % of the exposed persons. The quantum concept as well as equation (3) was combined by Riley [22] to equation (4).

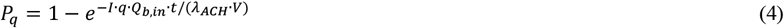

In equation (4), the relevant parameters are integrated. The probability of infection rises with the number of infectious persons (I), the quanta emission rate depending on the activity (q), the pulmonary ventilation rate of exposed susceptible persons (Q_b,in_), the duration of stay (t), but is inversely related to the air change rate (*λ*_*ACH*_) and the room volume (V).

Equation (5) can be used to calculate the individual risk of infection depending on the ratio of the number of inhaled viral copies N and the number of viral copies necessary to result in an infection N_0_ [24].

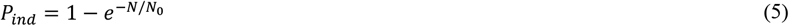

The probability P in equations (2) to (5) can be seen as the individual risk of infection P_ind_. If it is assumed, that statistically in a group of susceptible persons (N_Pers_) exactly this percentage of people is getting infected we define the attack rate (AR) in a given situation as the situational Predicted Attack Rate PAR_S_ (see equation (6)).

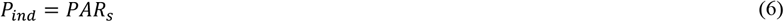

In poorly ventilated rooms, the assumption of a steady concentration of virus copies is often not fulfilled. The normalized time-dependent concentration process can be calculated according to equation (7) and is shown in Figure 1, assuming that the particles are immediately distributed uniformly in the room [25]. How rapidly the concentration of a person emitted contamination in a room rises depends on the overall lambda (λ_g_) and the time (t). Overall lambda thereby consists of the air change rate as well as the decay rates because of sedimentation and inactivation. The relative concentration (c_rel_) based on the steady-state concentration can be seen as an increase in the concentration compared to the volume flow.

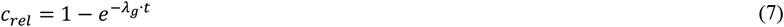

**Figure 1:**
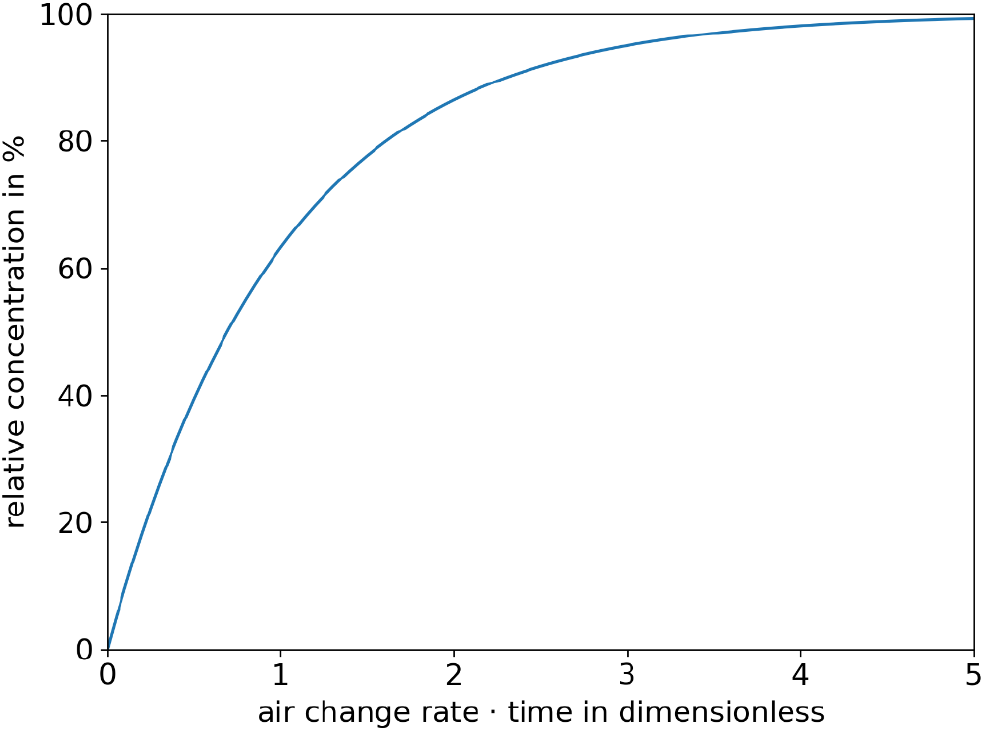
Relative concentration curve as a function of air change rate and time, based on the steady state concentration.

In many published studies that use the Wells-Riley equation to determine infection risk ideal mixing ventilation was assumed, which means that particles are evenly distributed in the room air immediately after emission. It should be considered that local concentration differences will occur in the room and that the viral load in the room areas will vary widely [26, 27, 28]. To estimate the risk of infection in a given setting by a given infectious person with the Wells-Riley-equation the quanta emission rate (q) has to be known. In various studies of infection occurrences associated with SARS-CoV-2, q was determined using the Wells-Riley equation retrospectively. Different authors [29, 30] found a range of 36 to 62 quanta/h with an assumed low activity (breathing, speaking) and values of 341 to 1190 quanta/h when singing. Furthermore, Buonanno et al. [31] as well as Bazant [32] used the viral load measured in the sputum of infected persons to calculate q for different activities. Therefore, emission rates for different activities (breathing volume flows, particle emission) as well as different states of infection (viral load in the sputum) can be calculated. A model, which applies this approach was set up by Lelieveld et al. [33].

In measurements of different research groups [34, 35, 36, 37, 38] the particle emission rates during breathing, speaking, coughing as well as singing were measured. During breathing through the nose between 25 particles/s [35, 37] and 135 particles/s [36] was emitted and during coughing about 13,700 particles/cough [35], whereas it can be seen that depending on the activity a wide range of particle emission rates can be found. The particle emissions while speaking and singing depended on the loudness of the activity, but in most cases the emission rates was found to be higher for singing than for speaking. Whereas for normal speaking the emission rates ranged between 30 particles/s [37] and 270 particles/s [36, 35], for singing it ranged approximately between 100 particles/s [37] and 2000 particles/s [34, 39].

In all five studies [34, 35, 36, 37, 39] regarding the particle emission rate of adults at least 99 % of the measured particles were smaller than 3.0 μm. In Alsved et al. [36] and Gregson et al. [37] 60 % of the particles were even smaller than 1.0 μm, whereas in Hartmann et al. [35] as well as Mürbe et al. [34] even 85 % of the particles were smaller than 1.0 μm and 60 % smaller than 0.5 μm. The particle emission rates measured by some of the authors are shown in Figure 2 and Table 2 (in the Appendix).

**Figure 2:**
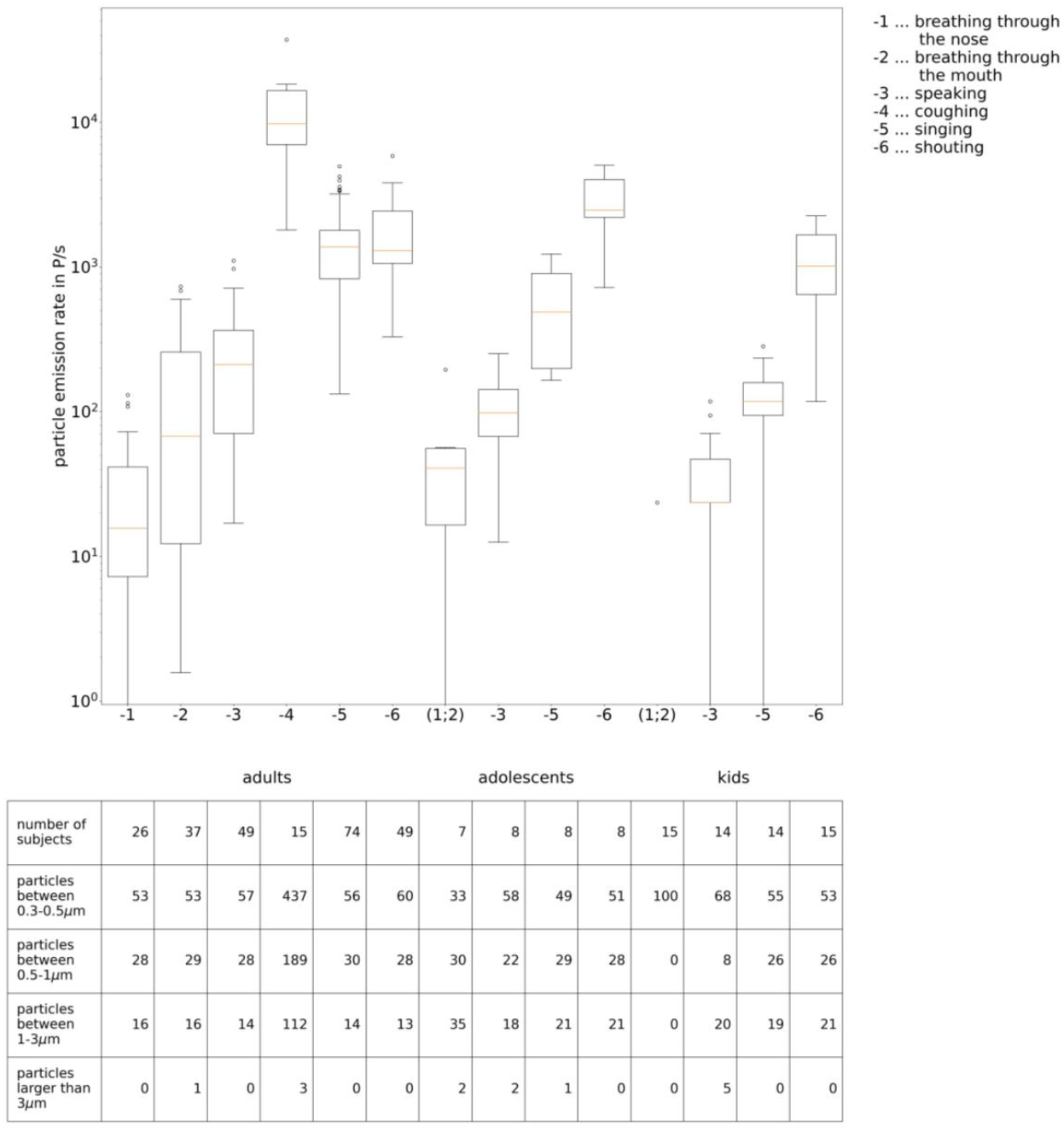
particle emission rates measured by some of the authors, for adults: [31, 36, 32], for adolescents [35] and for children [38]

Smaller particles remain airborne for a long time. The viral load (copies/ml) is aerosolized in the respiratory tract and not every small particle carries one virus. The larger the particle size, the higher the number of particles that actually carry a virus. Nevertheless, the measured particle size is the equilibrium size after evaporation. In different studies, a particle reduction to between 33 and 50% [40] of the original size was measured.

Face masks can be seen as a measure to reduce the number of emitted particles as well as the inhalation of particles from the air. The efficiency of a face mask thereby depends on three aspects:

- the filter efficiency of the fabric
- the leakage (i.e. air flow bypassing the mask) during exhalation
- the leakage during inhalation

Whereas the filter efficiency of medical masks, like surgical masks, FFP2-masks, N95 or KN95 is regulated, the efficiency of cotton masks or other homemade masks can vary widely. The results of the investigation of the filter efficiency of different materials were reviewed by Kwong et al. [41]. For example, for microfiber the filter efficiency ranged from 10 to 75% and for cotton/synthetic mix from 5 to 45%. The authors discuss that the materials have to be described with more details to make it possible to compare different studies.

Karuppasamy and Obuchowski [42] as well as Mueller et al. [43] investigated the influence if masks worn more tightly to the face. Whereas Karuppasamy and Obuchowski [42] found an improvement with surgical face masks, fixed with medical tape to the face of health care workers, Mueller et al. [43] found a reduced emission if the masks are tighter by using a nylon stocking to fix a cotton mask. The research group of Asadi et al. [44] and Cappa et al. [45] investigated the particle emission rate for different activities as well as different mask types. They found that the overall particle emission rate (through the mask as well as through leakages) is about 90% lower for coughing and 70% lower for talking compared to the case without masks. Still, this cannot be applied for cotton masks, where sometimes higher particle emission rates were measured with mask than without mask. In case of a reduction, especially larger particles were found to be reduced.

In a study by some of the authors [46] the ratio of air leaking around the edges of the masks while exhaling was measured. The leakage ranged between 20 and 90% for cotton masks, between 35 and 90% for surgical masks and between 5 and 75% for FFP1-masks. The results are comparable to results of Dreller et al. [47], who measured the leakage while inhaling. Nevertheless, in a study of Ueki et al. [48] the influence of a mask on a mannequin emitting virus laden particles was higher than a mask on the receiver on the amount of virus measured as an inhalation of the receiver. The difference can be explained by the differences in airflow. Whereas for exhaling especially large particles cannot follow the airflow and will be separated at the mask, for inhalation just smaller particles are still in the air. Therefore, the leakage might be the same, but the number of bypassing viruses is different between exhalation and inhalation, whereas masks are more helpful for the emitter than for the receiver.

Besides the number of emitted pathogen-laden aerosol particles, the number of inhaled pathogens is playing an important role as well with regard to the assessment of the risk of infection. The pulmonary ventilation rate may differ with different activities. Gupta et al. [49] performed a study with 25 healthy adults and found a sine wave for mere breathing, but a more constant volume flow during talking. In measurements with athletes as well as sedentary persons a maximum volume flow for the athletes of 200 l/min (12 m^3^/h) was found by Córdova and Latasa [50]. To measure the airflow without movement restrictions, a helmet was used by Jiang et al. [51] in 32 subjects (16 males, 16 females) during speaking with different volumes as well as during singing.

A comparison between a machine-learning based model and measurements of respiratory rate was performed by Dumond et al. [52].

As a conclusion, the following average values can be used for adults:

- low activity (breathing while lying): 0.45 m^3^/h [51]
- low activity (breathing while sitting, standing or talking): 0.54 m^3^/h [51, 52]
- singing: 0.65 m^3^/h [53]
- mid activity (physical work): 0.9 m^3^/h [52]
- sports: 1.2 m^3^/h [50, 52]

For children, the lung volume is smaller. Therefore, the respiratory rate for children aged 14 years can be assumed to be 0.45 m^3^/h for low activity (breathing while sitting, standing, talking) [54].

Antigen tests are currently widely used to detect infected individuals. The sensitivity depends on the quality of the used product and the viral load of the tested person. This was also discussed by some authors considering the quite different sensitivities of the rapid antigen tests performed by professionals (40 % [55], 60.9 % [56], 64.4 % [57], 64.5 % [56], 79.5 % [58] or 85 % [59]) as well as self-tests (74.4 % [58] and 82.5 % [59]). In a technical report of the British Department for Health and Social Care [60] the sensitivity of rapid tests depending on the viral load is given as 96 % for more than 1E+07 viral copies/ml, 92 % for 1E+04 to 1E+07 viral copies/ml and 43 % for lower viral loads. The value for the highest viral load was also confirmed by e.g. Lindner et al. [59], but seems pretty high for the other groups compared to the values found in the aforementioned studies. In most cases, where the infected person was not detected with the rapid test, the viral load was lower than 1E+06 viral copies/ml. In [61] it was shown that suitable test kits have a sensitivity of 80% compared with RT-PCR at a viral load of 1E+06 viral copies/ml. Even the lowest sensitive test showed a 90 % probable detection rate at a viral load of 2.3E+07 viral copies/ml. Similar orders of magnitude were found in [62]. Of 122 rapid antigen tests investigated by Scheiblauer et al. [63] 96 passed a limit of 75 % sensitivity at a viral load of 1E+06 viral copies/ml. No significant change in the test sensitivity for the VOC was found [64, 65]. In a model [66] as well as a longitudinal study [67] it was shown, that rapid antigen tests are able to detect infected persons during the course of an infection and may therefore reduce the transmission [66] if performed at a regular frequency [67]. Whereas the viral load can increase by a factor of about 100 within 24 h before symptom onset/peak viral load [13]. Rapid antigen tests will detect an infection only within the diagnostic window around the highest peak of infection. It is therefore possible for an individual to receive a negative test result for a rapid antigen test despite being infected and even contagious for other persons. For this reason, an increase in regular testing frequency can greatly increase the significance of a negative test result of a rapid antigen test compared with a negative result obtained with sporadic testing. It was shown in [68], that students who had close contact with a classmate who tested positive for SARS-CoV-2 and were subsequently tested daily avoided days absent from school, with no impact on overall infection incidence.

## Objectives

For the current study, the following research questions are derived:

- Which viral loads are necessary to infect others via aerosol?
- Which are the most influencing factors regarding airborne transmission?
- Can a risk assessment model be simplified to allow practical recommendations?
- Is there a possibility to implement a simple measurement system for infection risk?
- What is the impact of different prevention measures on the risk of airborne transmission?

## Methods

### Dose-Response Model to predict the individual infection risk and the Predicted Attack Rate (PAR)

Equations (4) and (5) assume an immediate homogeneous distribution of all emitted respiratory viruses within the room and steady state (time independent) situations.

In the following consideration, two different cases, one stationary and one time-dependent, are taken into account. For unsteady situations, it is assumed that the infected person enters the room at time 0 and the concentration in the room increases until a steady state is reached (see Figure 1).

The number of inhaled particles N_inh_ in equation (5) can be described with the help of equation (8). S_V_ is thereby the viral-emission rate in viral copies/time and 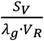 is the viral concentration per cubic meter of air. The overall lambda consists of the air change rate (ACH = *λ*_ACH_), the decay rate because of inactivation (*λ*_in_) and the decay rate because of sedimentation (*λ*_sed_).

Finally, equation (11) can be set up.

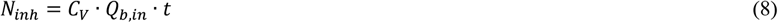

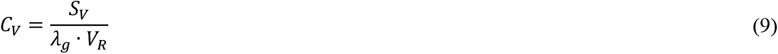

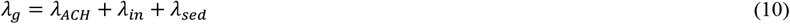

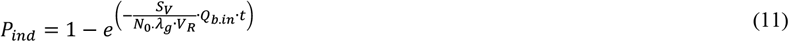

If face masks are used, the number of inhaled particles N_inh_ can be reduced. This reduction can be implemented into equation (11) as factor f_M_, which will result in equation (12).

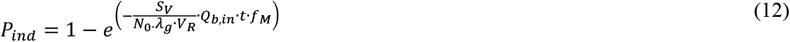

For the unsteady calculation, the course of the virus concentration is used according to equation (7). Equation (5) therefore transforms into equation (13) and equation (4) into equation (14).

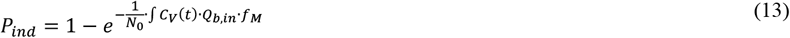

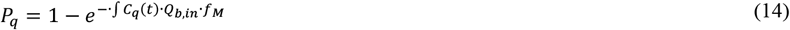

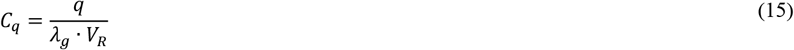

If the individual infection risk P_ind_ approximates statistically to P_q_ (equation (16)) equation (17) can be received, where PAR_S_ is defined as the situational Predicted Attack Rate, the Attack Rate during the stay in a room with infected persons.

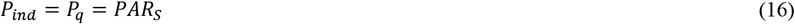

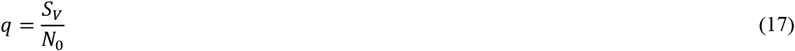

To retrospectively analyze the outbreaks, investigated in the following considerations, four categories of influencing factors are distinguished: first the emission rate (viral copies per time) in connection with the critical number of virus to result in an infection N_0_, second the parameter C_R_, which takes the boundary conditions of the room as well as the time of stay into account, third, the breathing volume flow of the inhaling person Q_b,in_ and fourth the total filter efficiency of the face masks considered by the filter factor f_M_.

#### 1. Virus related factor (VF)

The emission rate of virus laden particles depends on the activity, which influences the number of emitted particles as well as their size distribution. Furthermore, the viral load influences the number of virus carried on one particle. The emission rate S_v_ can therefore be described as the product of the particle emission rate *N*_*p*_, a factor considering their size distribution f_p_ and the viral load n_v_ (see equation (18)).

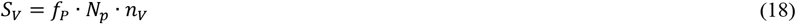

Thereby f_p_ describes a conversion factor from the particle emission rate per second to their volume emission rate per hour and depends on the size distribution. In the following calculations a value of 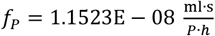 is used. The calculation of this conversion factor is displayed in the appendix.

N_0_ is assumed to be in the range of 100 to 300 viral copies [24]. The virus related factor is defined in equation (17).

#### 2. Situation-related factor (SF)

In the situational factor the boundary conditions for the specific situation are considered. It therefore consists of the room volume V_R_, the overall lambda (*λ*_*g*_) as well as the time of stay (t). In a steady state it can therefore easily be derived from equation (12) and will result in equation (19). For an unsteady situation the equation for the concentration (see Figure 1 and equation (20)) has to be integrated to get equation (21).

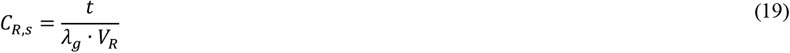

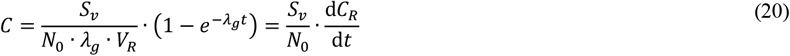

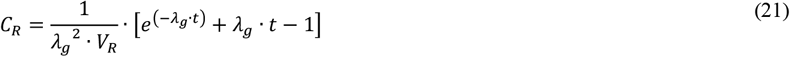

#### 3. Susceptible person-related factor (SPF)

As mentioned in the state of the arts the breathing volume flow depends on the activity of the persons. Furthermore, it can be split up into the exhalation flow rate (Q_b,ex_) of the infected person and the inhalation flow rate (Q_b,in_) of the susceptible persons. To calculate the number of inhaled particles, just the inhaled volume flow rate (Q_b,in_) has to be considered.

#### 4. Personal protection measures related factor (PPF)

To calculate the total efficiency of a face mask different factors for the infected person and the susceptible persons has to be considered.

Whereas the efficiency of the face mask carried by the infected person (*f*_*m,e*_) is characterized by the reduction of the number of virus laden aerosol particles introduced into the room air, the efficiency of the mask carried by the susceptible persons is characterized by their ability to reduce the number of inhaled virus laden aerosol particles (*f*_*m,in*_). To take into account that these factors are different equation (22) considers the total efficiency as the product of the two efficiencies, whereas f_m,e_ as well as f_m,in_ is the ratio of particles going by the mask or the difference between 1 and the ratio of particles separated by the mask.

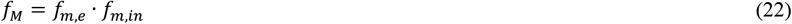

Equation (13) can therefore also be expressed as:

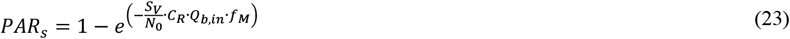

For the calculation of PARs the following assumptions must be considered:

- the aerosol is ideally mixed in the room
- the near field can contain a much higher virus-laden particle concentration, but it is neglected in the following
- the air, which is introduced into the room, is free of virus-laden particles
- a constant decay rate of deposition occurs (in this consideration 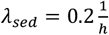)
- a constant decay rate because of inactivation occurs (in this investigation 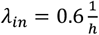)
- the concentration of virus-laden particles at the beginning of unsteady cases is 0 virus-laden particles/m^3^

Twenty-five different outbreaks either scientifically published or registered by the local health authorities were selected. Just publications considering the time of stay, the activity of the persons as well as the room conditions (size as well as ventilation) are taken into consideration. In addition, the outbreaks either had to be attributed to the wild-type of SARS-CoV-2 (e.g. by sequencing) or had occurred before 01.01.2021. Smaller outbreaks provided by local health authorities are included if they meet the same criteria. The description of the outbreaks as well as the boundary conditions for the calculations of these situations can be found in the appendix and in Table 3. The values were either taken from the publication or calculated from data given in them. Some data is due to better documentation more secured, whereas other values are somewhat less certain, a variance of the values is assumed. A normal distribution of the values characterized by the mean value and the standard deviation (given as the variance) was assumed. A Monte-Carlo-Simulation was used to randomly combine values from within this range for the calculation of the outbreaks [69]. Therefore, a conclusion can be drawn, about the reliability of the calculated values and their variance evoked by the uncertain boundary conditions. The outbreaks can be separated into different categories: choir rehearsals (4 outbreaks), outbreaks with higher physical activity (4 outbreaks), meetings with lower activity, but many people (3 outbreaks), outbreaks in public transport (6 outbreaks) and smaller outbreaks, which are sometimes not scientifically published, but investigated by local health authorities (8 outbreaks).

## Results

For the investigated outbreaks and their known boundary conditions the virus-related factor (*S*_*V*_/*N*_0_) was calculated retrospectively. Besides the results for the virus-related factor also the intermediate results for the other factors C_R_ (steady (regarding equation (19)) and unsteady (regarding equation (21))), f_M_ and O_b,in_ can be seen in Table 1.

**Table 1:**
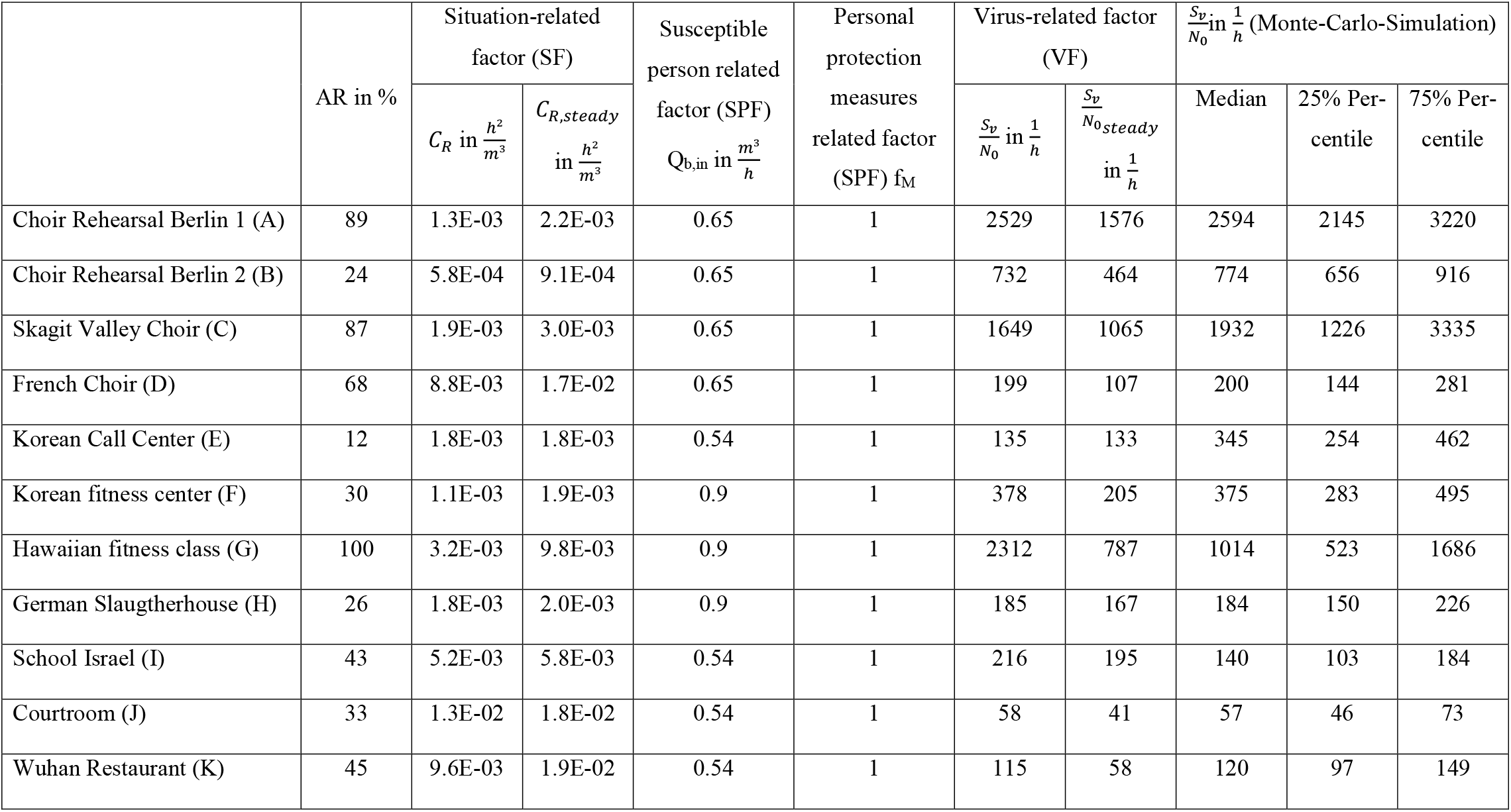
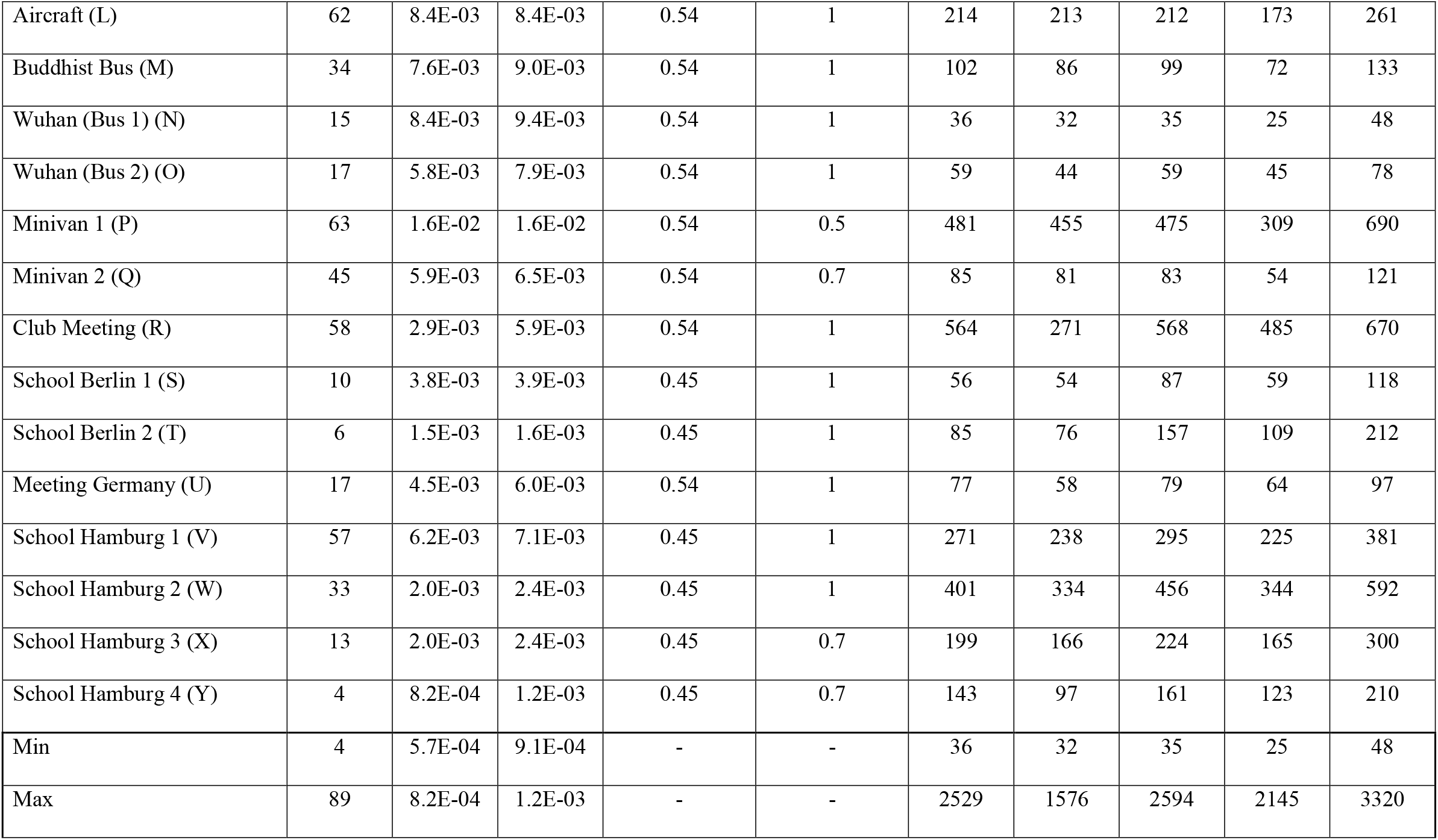
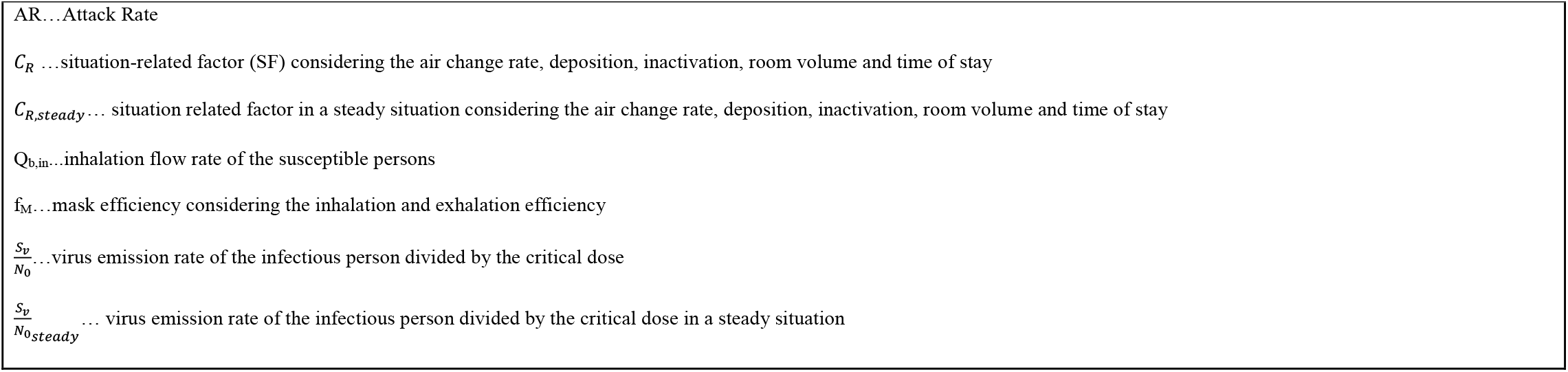
Results for the different factors for the investigated outbreaks

**Table 2:**
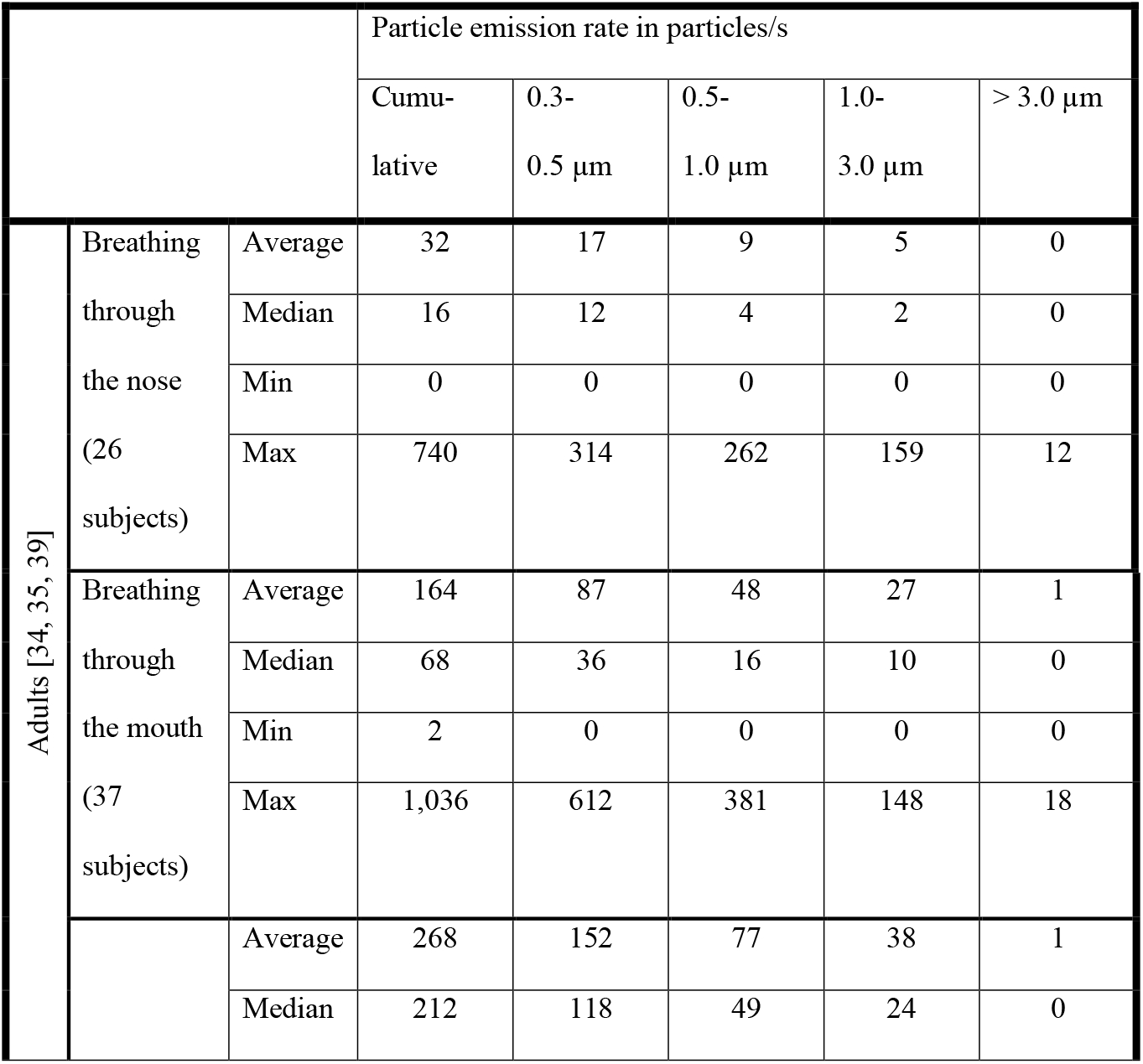
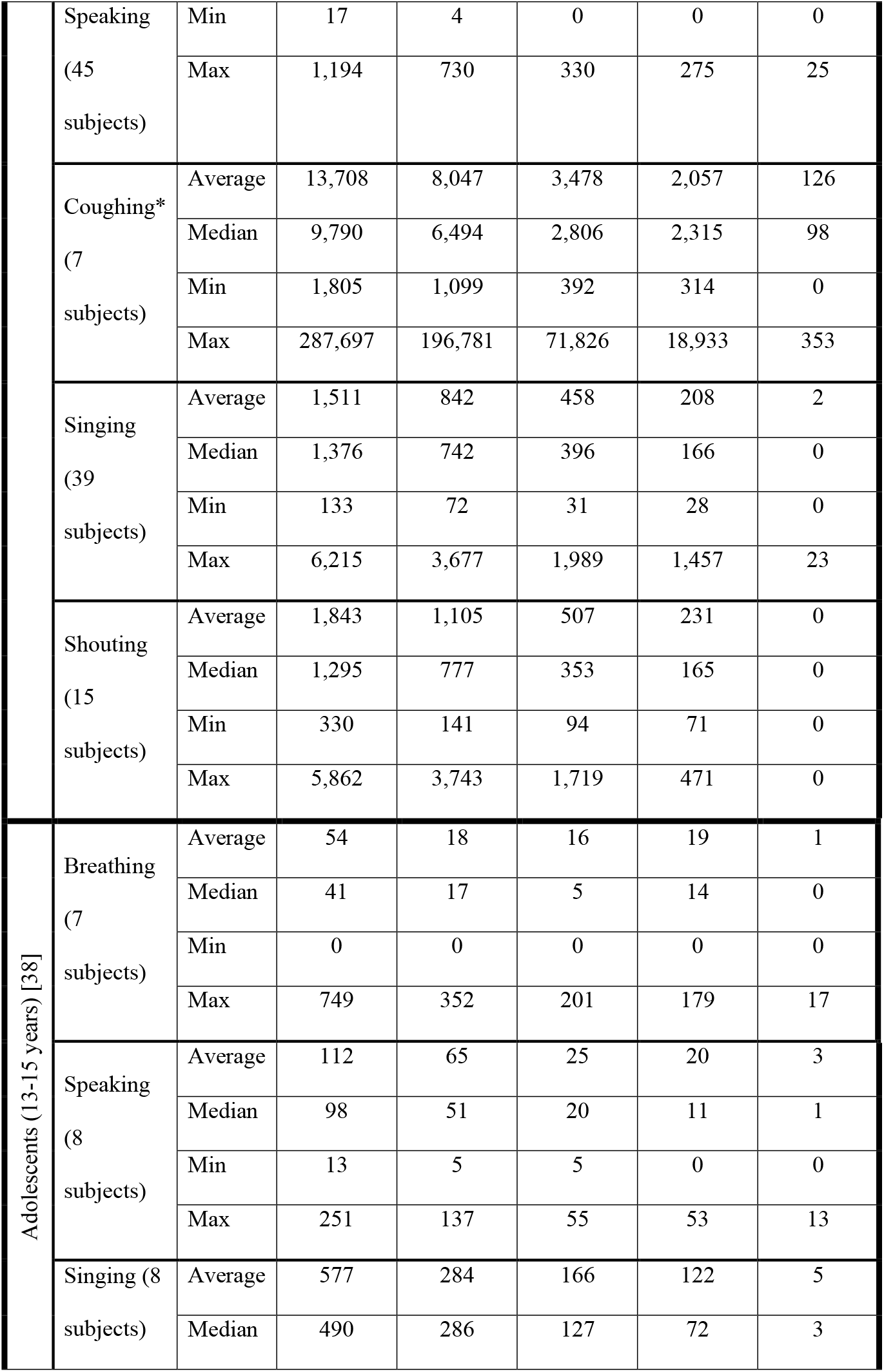
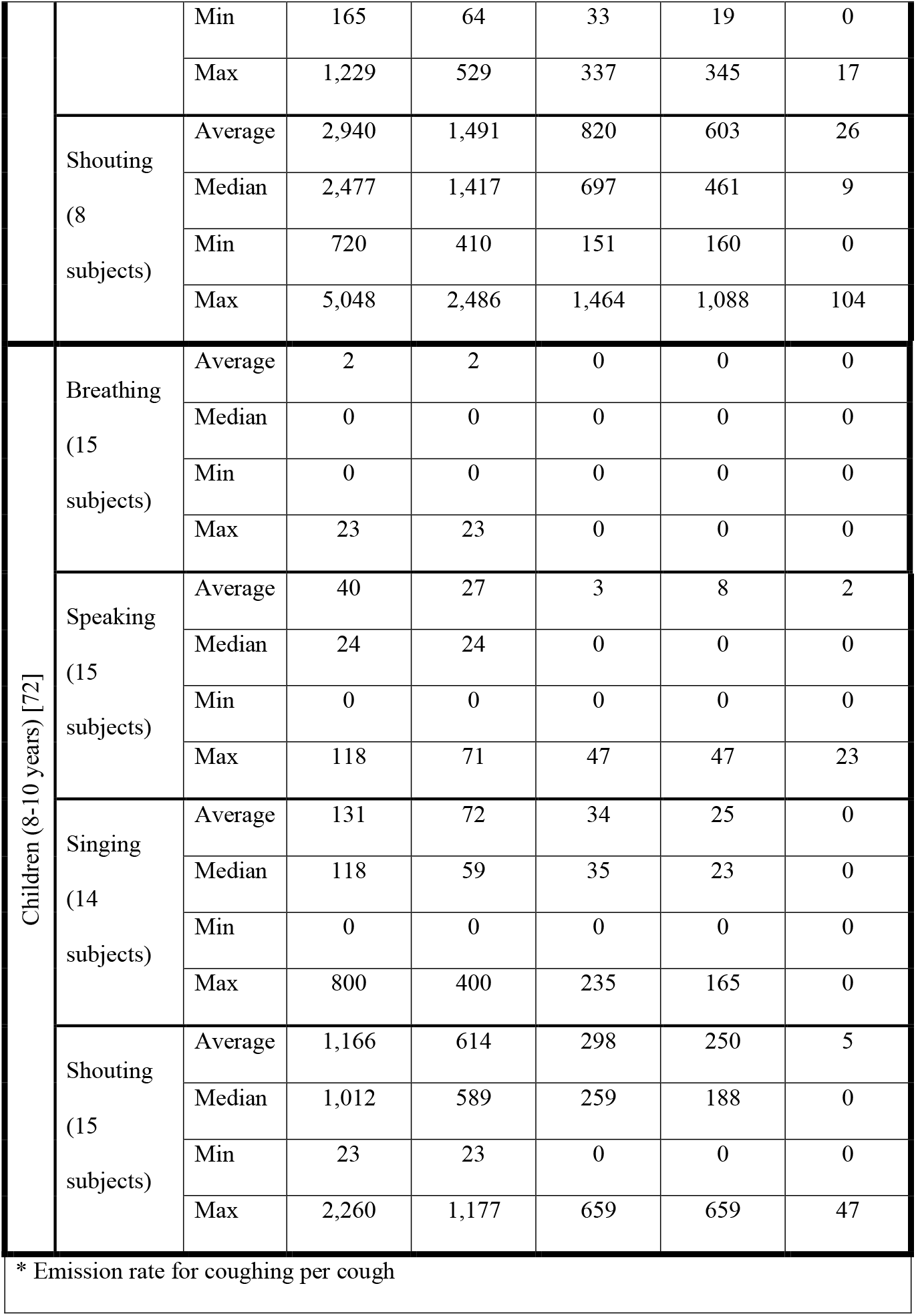
Particle emission rates measured by the authors, partially published for adults in [34, 39, 35], for adolescents in [38] and for children in [72]

We compared the results from the retrospective investigation with available data regarding the viral load and the particle emission. Therefore, in Figure 3 and Figure 4 the ratio of the viral emission and the critical dose (*S*_*V*_/*N*_0_) are presented over the viral load and the particle emission. The colors visualize the AR from the investigated outbreaks, whereby the single outbreaks are shown as dots. A mean particle emission rate as a function of activity was assumed when plotting the dots, according to Figure 2. In Figure 3 the critical dose N_0_ is assumed to be the minimal value of 100 viral copies and in Figure 4 a higher value of 300 viral copies, both related to [24]. With a higher critical dose, the lines with similar PARs are shifted upwards, whereas either a higher viral load or a higher particle emission rate is necessary to result in the same PARs. It can be seen that the viral load for all investigated outbreaks had to be higher than 1E+08 viral copies/ml to explain the outbreaks with the given boundary conditions. If instead of equation (21) for time-depended calculation, equation (19) for steady-state assumption is used, the values 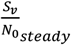 are lower than the values calculated for the unsteady conditions. Nevertheless, the viral load had to be higher than 1E+08 viral copies/ml to reach the ARs (see Table 1).

**Figure 3:**
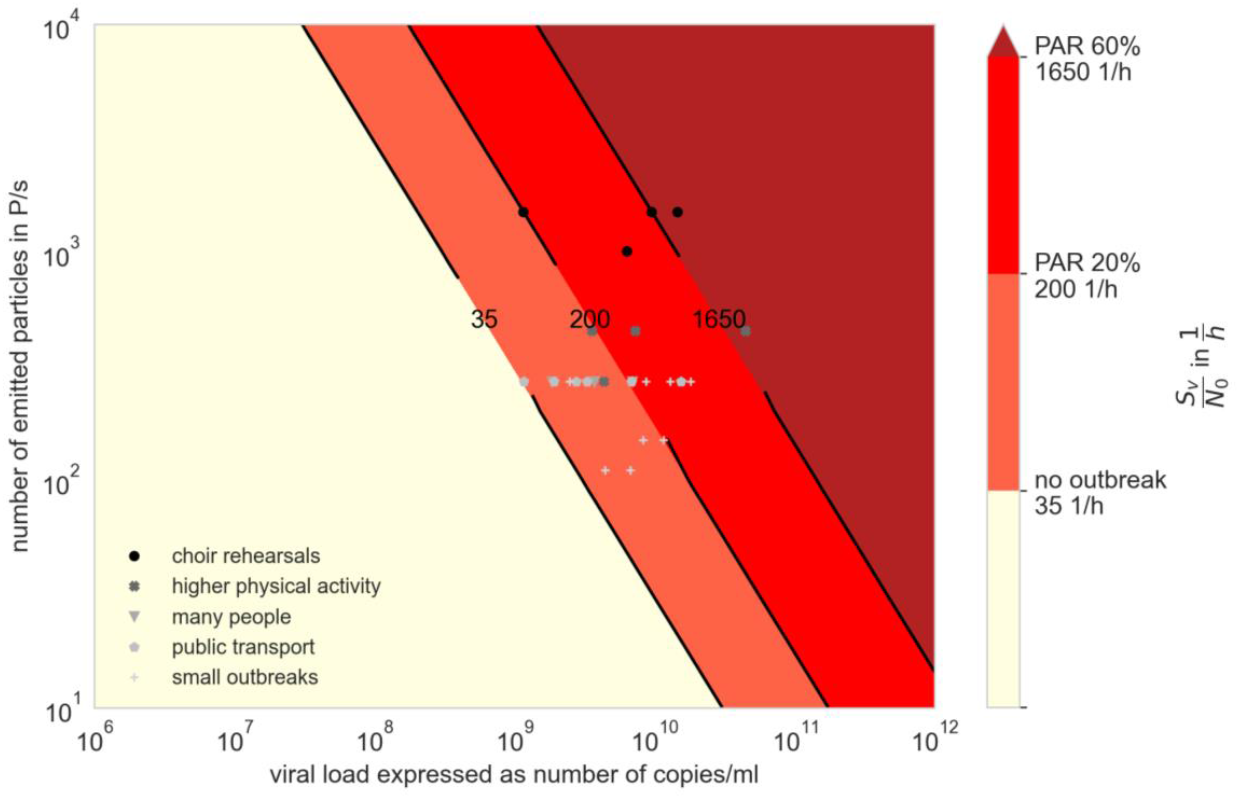
Virus Factor (VF) for different viral loads and particle emission rates with N_0_=100 viral copies, the attack rates found in the investigated outbreaks are shown with the different colors; the markers show the amount of virus factor at assumed mean particle emission rate of the related activity according to Figure 2.

**Figure 4:**
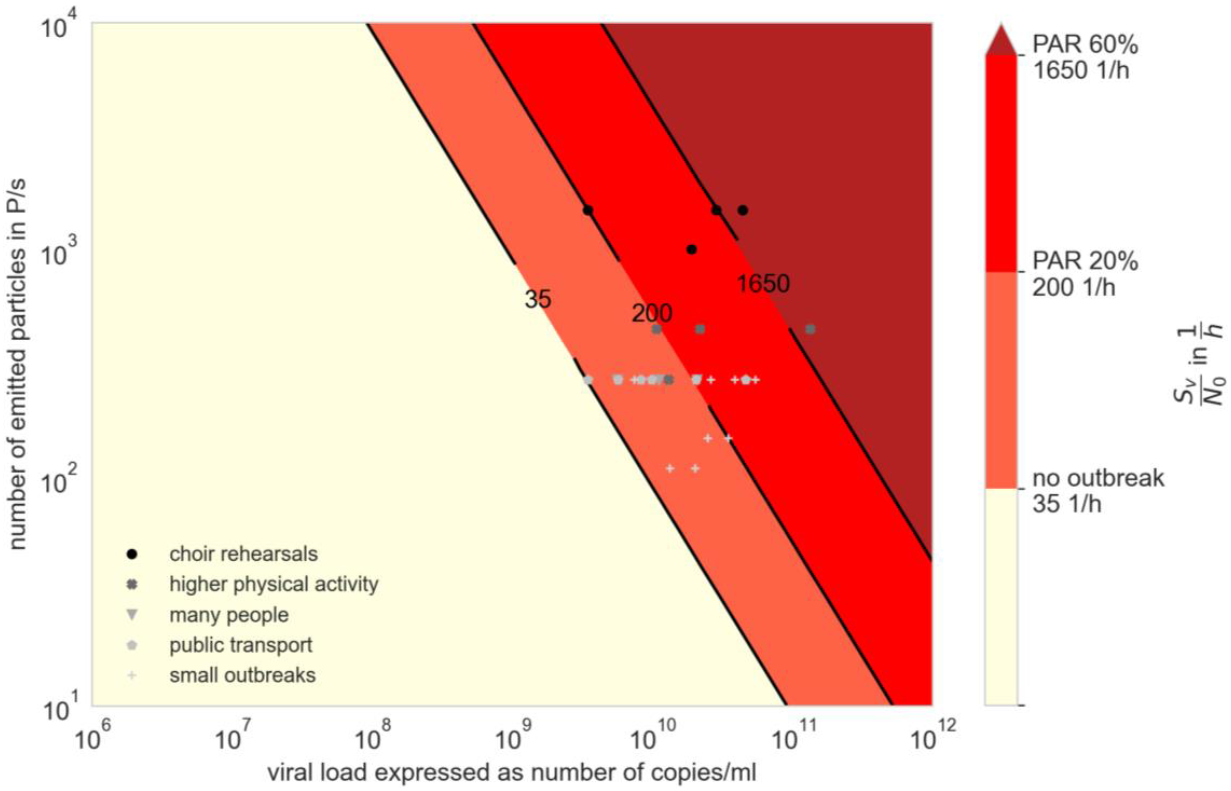
Virus Factor (VF) for different viral loads and particle emission rates with N_0_=300 viral copies, the attack rates found in the investigated outbreaks are shown with the different colors; the markers show the amount of virus factor at assumed mean particle emission rate of the related activity according to Figure 2.

As it is mentioned in Table 3 some of the boundary conditions have been assumed afterwards, so they are not as certain as other pieces of information. The certainty of the different boundary conditions was evaluated as quite secure (standard deviation ± 5 %), a bit insecure (standard deviation ± 20 %) and insecure or unknown (standard deviation ± 50 %) for each outbreak as it is displayed in Table 3 in red. To take these uncertainties into consideration a Monte-Carlo-Simulation with 10,000 simulations for each outbreak is performed. The investigated aspects (air change rate, room volume, number of infected persons, if it cannot be secured, that it was just one person, breathing volume flow, time of stay and attack rate) are assumed to be normally distributed with the given value as mean and the assumed level of security as standard deviation. Furthermore, limits for the AR, lower than 100% and the other aspects larger than 0, are considered. The median as well as the 25 %-percentile and the 75 %-percentile are displayed in Table 1.

**Table 3:** scientifically published SARS-CoV-2 outbreaks small outbreaks investigated by Robert-Koch-Institute or local health authorities

|  | Choir Rehearsal Berlin 1 | Choir Rehearsal Berlin 2 | Skagit Valley Choir | French Choir | Korean Call Center | Korean fitness center | Hawaiian fitness class | German Slaught-house |
| --- | --- | --- | --- | --- | --- | --- | --- | --- |
| Source/s | [39] | [39] | [30, 29] | [73] | [30, 76] | [30, 77] | [74] | [75] |
| Air change rate in 1/h | 0.17 | 0.45 | [30] 1.5, [29] 0.3-1.0 | unventilated, assumed 0.1 | [30] 1.5 | [30] 1.5 | unventilated, assumed 0.1 | 0.53 |
| | 0 $\pm 50\%$ | 0 $\pm 50\%$ | 0 $\pm 50\%$ | 0 $\pm 50\%$ | 0 $\pm 50\%$ | 0 $\pm 50\%$ | 0 $\pm 50\%$ | ++ $\pm 5\%$ |
| Room volume in m<sup>3</sup> | 1200 | 1720 | 810 | 135 | 1143** | 180 | 114*** | 3000 |
| | ++ $\pm 5\%$ | ++ $\pm 5\%$ | + $\pm 20\%$ | + $\pm 20\%$ | + $\pm 20\%$ | + $\pm 20\%$ | 0 $\pm 50\%$ | + $\pm 20\%$ |
| Number of previously infected persons | 1 | 1 | 1 | 1* | 1 | 1 | 1 | 1 |
| | ++ $\pm 5\%$ | ++ $\pm 5\%$ | ++ $\pm 5\%$ | 0 $\pm 50\%$ | ++ $\pm 5\%$ | ++ $\pm 5\%$ | ++ $\pm 5\%$ | ++ $\pm 5\%$ |
| Number of susceptible persons | 77 | 42 | 61 | 25 | 89 | 192 | 10 | 78 |
| Main activity | Singing | Singing/ Speaking | Singing | Singing | Speaking | Physical Activity | Physical Activity | Heavy working |
| Breathing volume flow ($Q_{b,e}$ and $Q_{b,in}$) in m<sup>3</sup>/h | 0.65 | 0.61 | 0.65 | 0.65 | 0.54 | 0.9 | 0.9 | 0.9 |
| | + $\pm 20\%$ | + $\pm 20\%$ | + $\pm 20\%$ | + $\pm 20\%$ | + $\pm 20\%$ | + $\pm 20\%$ | + $\pm 20\%$ | + $\pm 20\%$ |
| Mask efficiency in % | 1 | 1 | 1 | 1 | 1 | 1 | 1 | 1 |
|  | - | - | - | - | - | - | - | - |
| Time of stay in h | 2.5 | 2 | 2.5 | 2 | 8 | 0.8 | 1 | 8 |
| | ++ $\pm 5\%$ | ++ $\pm 5\%$ | ++ $\pm 5\%$ | ++ $\pm 5\%$ | + $\pm 20\%$ | ++ $\pm 5\%$ | ++ $\pm 5\%$ | + $\pm 20\%$ |
| Attack Rate in % | 89 | 24 | 87 | 68 | 65 | 30 | 100 | 26 |
| | ++ $\pm 5\%$ | ++ $\pm 5\%$ | + $\pm 20\%$ | + $\pm 20\%$ | ++ $\pm 5\%$ | + $\pm 20\%$ | + $\pm 20\%$ | ++ $\pm 5\%$ |
| * 3 persons had contact to a later on positive tested person, but just one person had symptom onset on the day after the rehearsal |  |  |  |  |  |  |  |  |
| ** transmission occurred just in a volume of 600 m<sup>3</sup> |  |  |  |  |  |  |  |  |
| *** area given, height of 3m assumed |  |  |  |  |  |  |  |  |
| Evaluation of the security of the boundary conditions ++ quite secure, + a bit insecure, 0 unknown/insecure, assumed level of deviation |  |  |  |  |  |  |  |  |

|  | School Israel | Courtroom | Wuhan Restaurant | Aircraft | Buddhist Bus | Wuhan (Bus 1) | Wuhan (Bus 2) | Minivan 1 | Minivan 2 |
| --- | --- | --- | --- | --- | --- | --- | --- | --- | --- |
| Source/s | [78] | [79] | [84, 1] | [80] | [30, 81] | [30, 82] | [30, 82] | [83] | [83] |
| Air change rate in 1/h | 2.7 | unknown, assumed 0.3 | [1] 0.56-0.77 | 21 | 3 | 3 | 3 | 9 | 9 |
| | 0 ±50% | 0 ±50% | 0 ±50% | + ±20% | 0 ±50% | 0 ±50% | 0 ±50% | 0 ±50% | 0 ±50% |
| Room volume in m<sup>3</sup> | 150 | 150 | [1] 431* | 60** | 50 | 71 | 34 | 16 | 16 |
| | ++ ±5% | ++ ±5% | + ±20% | ++ ±5% | + ±20% | + ±20% | + ±20% | + ±20% | + ±20% |
| Number of previously infected persons | 1 | 1 | 1 | 1 | 1 | 1 | 1 | 1 | 1 |
| | ++ ±5% | ++ ±5% | ++ ±5% | ++ ±5% | ++ ±5% | ++ ±5% | ++ ±5% | ++ ±5% | ++ ±5% |
| Number of susceptible persons | 66 | 9 | 21 | 19 | 68 | 48 | 12, 1 with mask | 4, all with cloth masks | 3, infected person w/o mask, other persons w/ mask |
| Main activity | Speaking | Speaking | Speaking | Speaking | Speaking | Speaking | Speaking | Speaking | Speaking |
| Breathing volume flow (Q<sub>b,e</sub> and Q<sub>b,in</sub>) in m<sup>3</sup>/h | 0.5 | 0.54 | 0.54 | 0.54 | 0.54 | 0.54 | 0.54 | 0.54 | 0.54 |
| | + ±20% | + ±20% | + ±20% | + ±20% | + ±20% | + ±20% | + ±20% | + ±20% | + ±20% |
| Mask efficiency in % | 1 | 1 | 1 | 1 | 1 | 1 | 1 | 0.7 | 0.85 |
| | - | - | - | - | - | - | - | + ±20% | + ±20% |
| Time of stay in h | 4.5 | 3 | 1.2 | 11 | 1.7 | 2.5 | 1.0 | 2 | 2 |
| | ++ ±5% | ++ ±5% | ++ ±5% | ++ ±5% | ++ ±5% | ++ ±5% | ++ ±5% | ++ ±5% | ++ ±5% |
| Attack Rate in % | 43 | 33 | 45 | 6 | 34 | 15 | 17 | 100 | 33 |
| | ++ ±5% | + ±20% | ++ ±5% | ++ ±5% | ++ ±5% | ++ ±5% | ++ ±5% | ++ ±5% | ++ ±5% |

\* Transmission occurred just in a volume of 47 m<sup>3</sup>
\*\* Only the Business Class is included where the majority of infections occurred

|  | Club Meeting | School Berlin 1 | School Berlin 2 | Meeting Germany | School Hamburg 1 | School Hamburg 2 | School Hamburg 3 | School Hamburg 4 |
| --- | --- | --- | --- | --- | --- | --- | --- | --- |
| Source/s | [85] | [85] | [85] | [85] | [86] | [86] | [86] | [86] |
| Air change rate in 1/h | 0.20<br>0 ±50% | 8.3<br>0 ±50% | 10<br>0 ±50% | 1.2<br>0 ±50% | 1.9<br>0 ±50% | 3.3<br>0 ±50% | 3.2<br>0 ±50% | 3.2<br>0 ±50% |
| Room volume in m<sup>3</sup> | 254<br>++ ±5% | 180<br>++ ±5% | 150<br>++ ±5% | 170<br>++ ±5% | 154<br>++ ±5% | 157<br>++ ±5% | 157<br>++ ±5% | 157<br>++ ±5% |
| Number of previously infected persons | 1<br>++ ±5% | 1<br>++ ±5% | 1<br>++ ±5% | 1<br>++ ±5% | 1<br>++ ±5% | 1<br>++ ±5% | 1<br>++ ±5% | 1<br>++ ±5% |
| Number of susceptible persons | 25 | 27 | 20 | 11 | 28 | 24 | 24 | 27 |
| Main activity | Speaking | Speaking | Speaking | Speaking | Speaking | Speaking | Speaking | Speaking |
| Breathing volume flow (Q<sub>b,e</sub> and Q<sub>b,in</sub>) in m<sup>3</sup>/h | 0.54<br>+ ±20% | 0.45<br>+ ±20% | 0.45<br>+ ±20% | 0.54<br>+ ±20% | 0.54/0.45*<br>+ ±20% | 0.54/0.45*<br>+ ±20% | 0.54/0.45*<br>+ ±20% | 0.54/0.45*<br>+ ±20% |
| Mask efficiency in % | 1<br>- | 1<br>- | 1<br>- | 1<br>- | 1<br>- | 1<br>- | 0.7<br>+ ±20% | 0.7<br>+ ±20% |
| Time of stay in h | 1.5<br>++ ±5% | 4.5<br>++ ±5% | 1.5<br>++ ±5% | 2<br>++ ±5% | 3<br>++ ±5% | 1.5<br>++ ±5% | 1.5<br>++ ±5% | 0.75<br>++ ±5% |
| Attack Rate in % | 58<br>++ ±5% | 10<br>++ ±5% | 6<br>++ ±5% | 17<br>++ ±5% | 57<br>++ ±5% | 33<br>++ ±5% | 13<br>++ ±5% | 4<br>++ ±5% |
| Evaluation of the security of the boundary conditions ++ quite secure, + a bit insecure, 0 unknown/insecure, assumed level of deviation<br>* exhalation/inhalation breathing volume flow |  |  |  |  |  |  |  |  |

In general, it can be seen, that the median agrees well with the values calculated from the most probable boundary conditions. The 25 % percentile is between 15 % and 48 % lower than the Median and the 75 %-percentile between 19 % and 72 % higher than the median. Cases with especially high deviation from the median are the Hawaiian fitness class and the Skagit Valley Choir, whereas most deviations ranged between 20 % and 35 %.

For the two choir outbreaks (Berlin 1 A and Skagit Valley C) with similar boundary conditions quite similar ratios of S_v_/N_0_ were calculated. For lower attack rates (French Choir, D) or less singing (Berlin 2, B) the emission rate was calculated to be lower and in the same range as for the call center I, the fitness classes (F, G) or the slaughterhouse (H). Furthermore, the outbreak in the restaurant (K), the Minivan 1 (P) and the Club Meeting (R) revealed such high values for the inhaled number of infectious particles compared to the critical dose. A possible explanation is that in these situations the infectious person talked louder, because other persons talked as well or the infectious person had a higher viral emission. For the outbreaks in schools (I, S, T, V, W, X, Y) the median values of S_v_/N_0_ ranged between 100 and 700 1/h, which is lower than for the choir or meeting outbreaks, but higher than for the other outbreaks correlated with public transport (L, M, N, O, Q).

### Derivation of simplified key figures and calculations for the assessment of infection risks and preventive measures

Equation (25) is created from equation (23) with the assumption (24), where R_S_ is defined as the situational reproduction number (the number of persons probably get infected during the situation), which should be valid statistically.

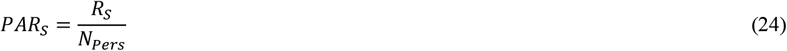

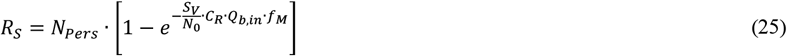

This equation can be transformed into equation (26).

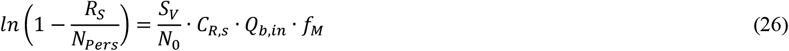

For the simplified calculation, a steady state situation is assumed. C_R,s_ can therefore be replaced by equation (19).

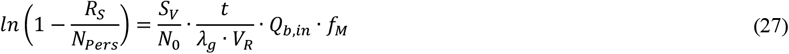

In the following, it is assumed that the air change rate is the dominant variable within equation (10). This is valid whenever non-residential standards and guidelines (DIN / ASHRAE) are respected. Therefore *λ*_*g*_ ≈ *λ*_*ACH*_ can be assumed. Taking equation (28) into account, equation (29) is obtained, with q_Pers_ the specific volume flow (person-related volume flow) in m^3^ per hour and person.

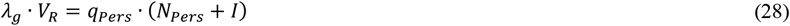

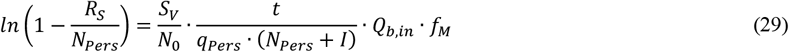

The following simplified method can easily be used in situations, where not more than one person shall be infected and therefore R_S_ = 1 and therefore no outbreak would probably happen due to aerosol transmission (***definition of outbreak***: more than one person get infected during transmission event). In rooms with number of susceptible persons *N*_*Pers*_ > 5 and *N*_*Pers*_ ≫ *I,* 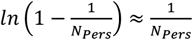 and 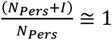 will result in a small error compared to the origin and equation (29) can be simplified into equation (30).

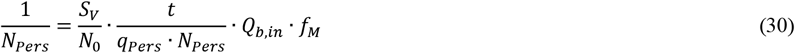

Further on, (30) can be converted into equation (31) to calculate the specific volume flow of virus free air *q*_*Pers*_ to fulfill R_S_ = 1.

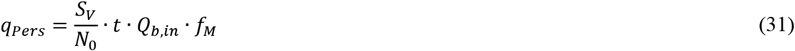

In case of supply of outdoor air as virus free volume flow *q*_*Pers*_ is correlated with a CO_2_-concentration. Whereas the number of inhaled particles increases linearly with time of stay (in case of steady state), the CO_2_-concentration does not change, caution has to be taken when using the CO_2_-concentration as indicator for a risk of infection.

Instead of a person related volume flow q_Pers_ the volume flow can also be calculated per hour time of stay (see equation (32)).

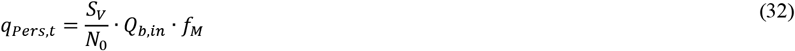

Figure 3 can be converted into Figure 5 for 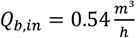 (low activity (breathing while sitting, standing or talking)). In Figure 5, the specific volume flow per person and h of stay necessary to infect not more than one further person is displayed.

**Figure 5:**
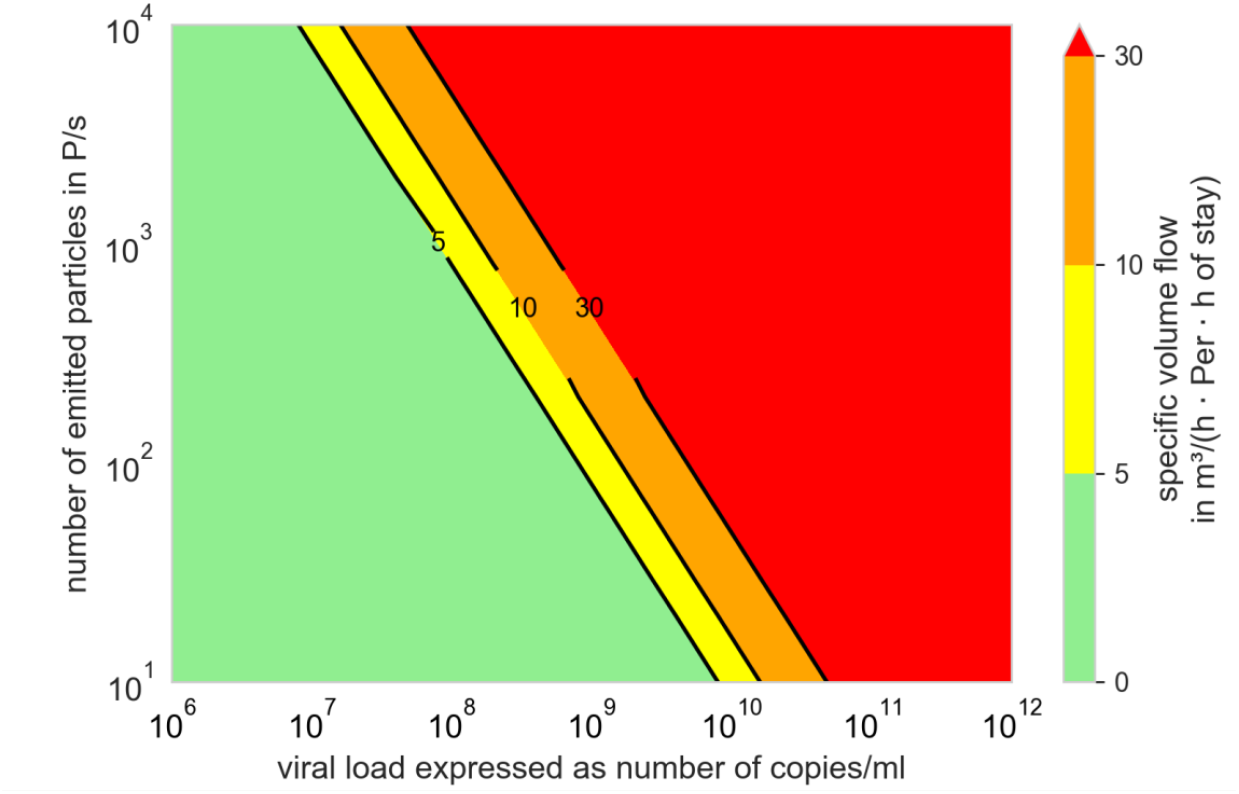
specific volume flow depending on the number of emitted, particles and the viral load to limit the number of newly infected persons to one, N_0_=100 viral copies, f_M_=1, Q_b,in_=Q_b,e_=0.54m^3^/h

The green marked area can easily be reached in most rooms for normal times of stay.

For the yellow and orange area short times of stay or further measures have to be considered to keep the number of newly infected persons below one (R_S_ ≤ 1), whereas a much higher air supply is necessary. The volume flows in the red area cannot be reached in rooms with common airflow rates.

Instead of a specific volume flow per person *q*_*Pers*_ a specific volume per person (V_Pers_) can be used together with the overall lambda (λ_g_) to convert equation (31) into equation (33).

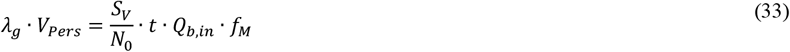

From this equation it can easily be seen that recommending an air change rate alone, e.g. 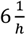 [70, 71], is not useful. In addition, the volume per person and the time of stay have to considered as well as virus-related properties and other preventive measures, e.g. wearing masks.

From equation (30), equation (34) can be derived. With this simplified approach also the maximal possible number of persons can be found, up to which not more than one further person will get infected in the specific situation. Therefore, the available volume flow Q has to be known.

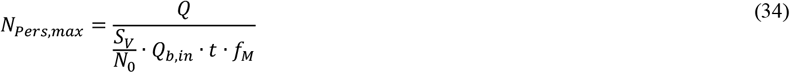

With the simplifications 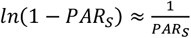 and 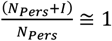, valid for *PAR*_*S*_ < 20% and *N*_*Pers*_ ≫ *I*, PAR_s_ according to equation (35) as well as R_s_ according to equation (36) can be predicted relatively good within the limited range of values.

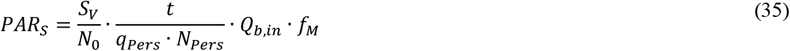

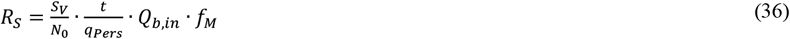

Equations (35) and (36) can be used to comparatively evaluate different situations in indoor environments as well as preventive measures. Therefore, a risk factor x_r_ can be defined according to equation (37). If the VF remains the same in the rooms being compared (identical virus variant), then the risk factor depends on SF, SPF, and PPF only.

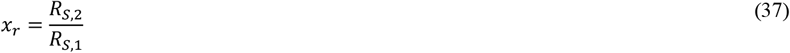

In Figure 6 same everyday life situations are compared to a 0.5 hour stay in a supermarket with mask, using equation (37). The details for these exemplary considerations can be found in the appendix, table 4.

**Table 4:**
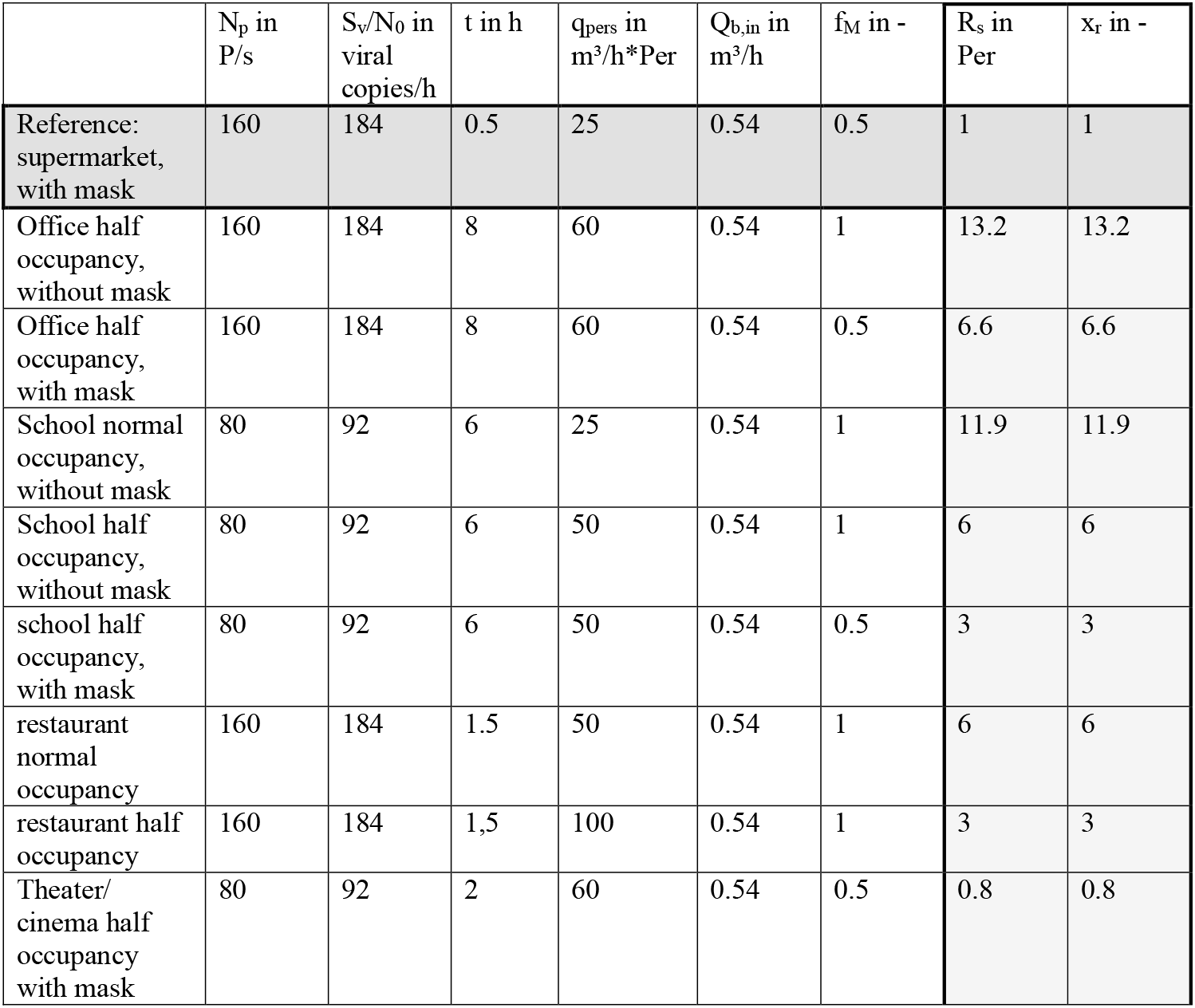
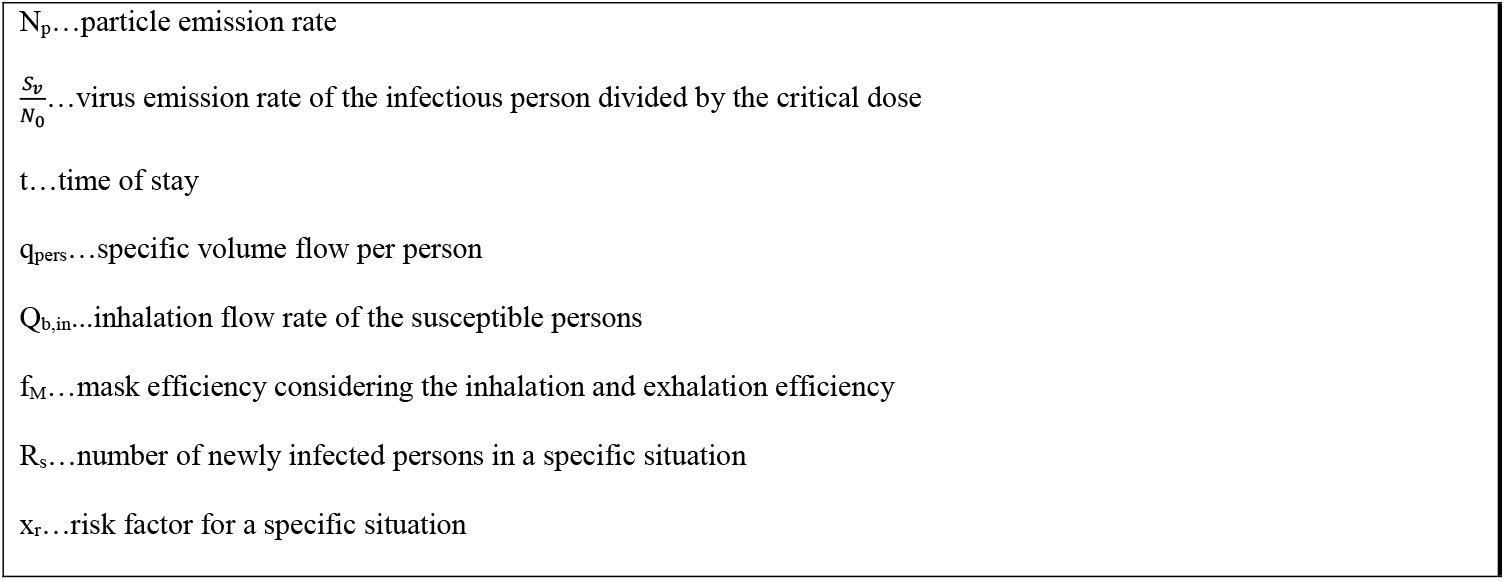
Comparison of different situations with the simplified approach

**Figure 6:**
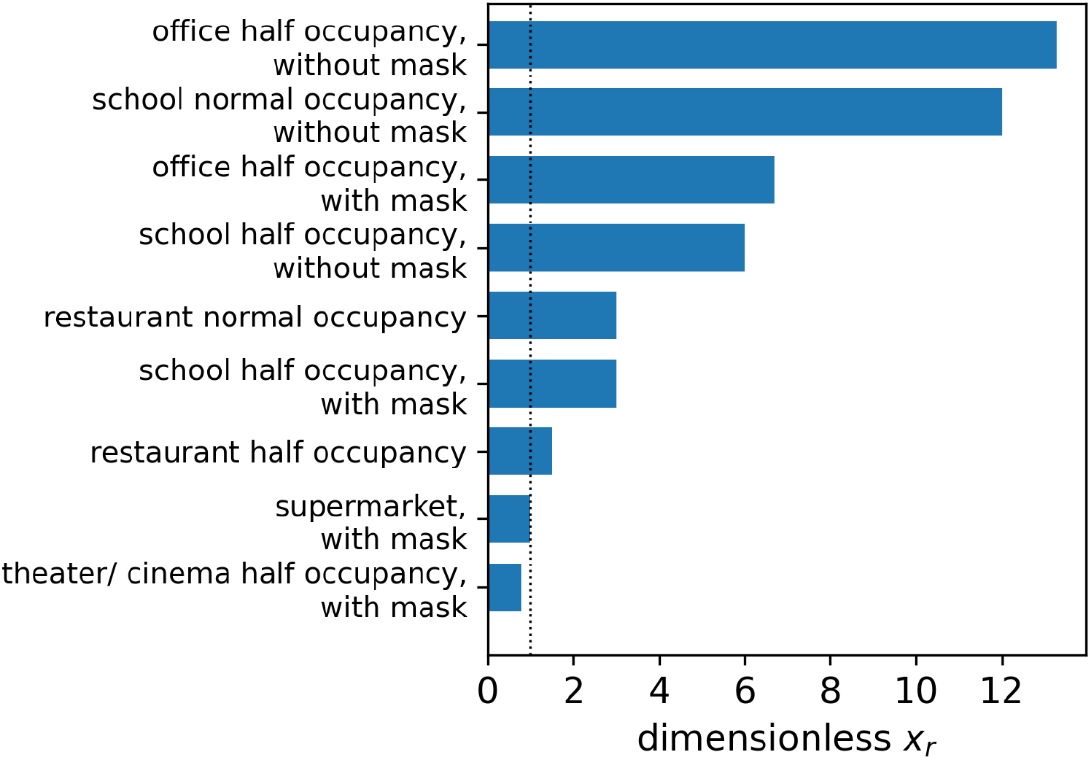
comparison of the risk factor x_r_ for different everyday life situations: o,5 hour stay in a supermarket, wearing a mask as reference

### Influence of Variants of concern (VOC)

For example, if the transmission rate doubles and this is not caused by a change in behavior (SF, SPF, or PPF), then it is due to the change in the VF. Here, either the necessary critical dose or the viral load or both may have shifted. The influence of the critical dose and S_v_/N_0_-ratio to the PAR_s_ is shown in Figure 7.

**Figure 7:**
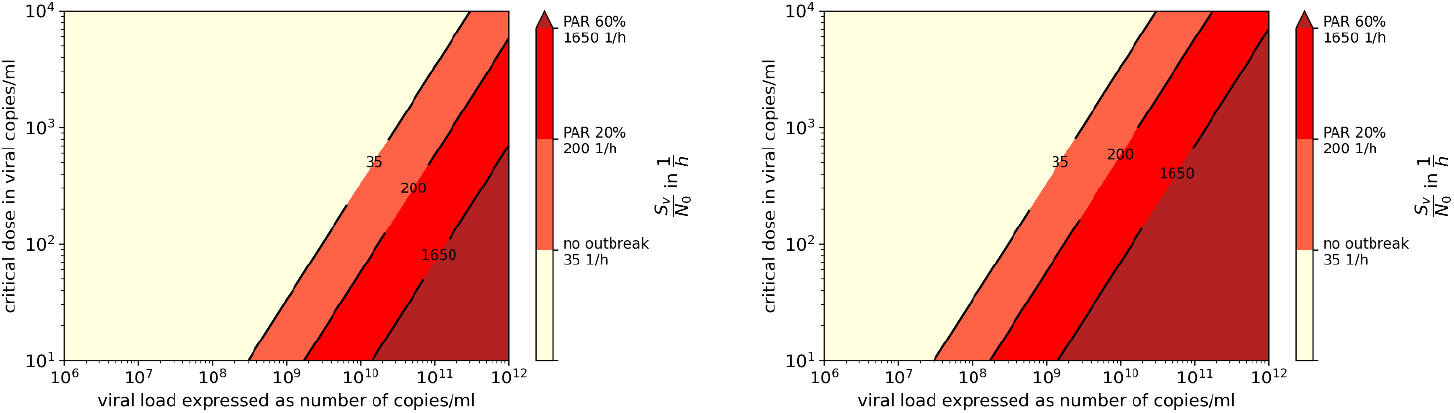
Virus Factor for different viral loads and critical doses with a particle emission rate of 100 P/s, (left) and 1,000 P/s (right), the attack rates found in the investigated outbreaks are shown with different colors

If for a first assumption it is assumed that the ratio of change in the transmissibility is correlated with the ratio of change in the R-value, which is furthermore inversely correlated with the critical dose it can be assumed that an increase of the transmissibility of 50 % 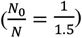 will result in a reduction of the critical dose from 100 viral copies to 67 viral copies. Figure 3 can therefore be converted into Figure 8 and Figure 5 into Figure 9.

**Figure 8:**
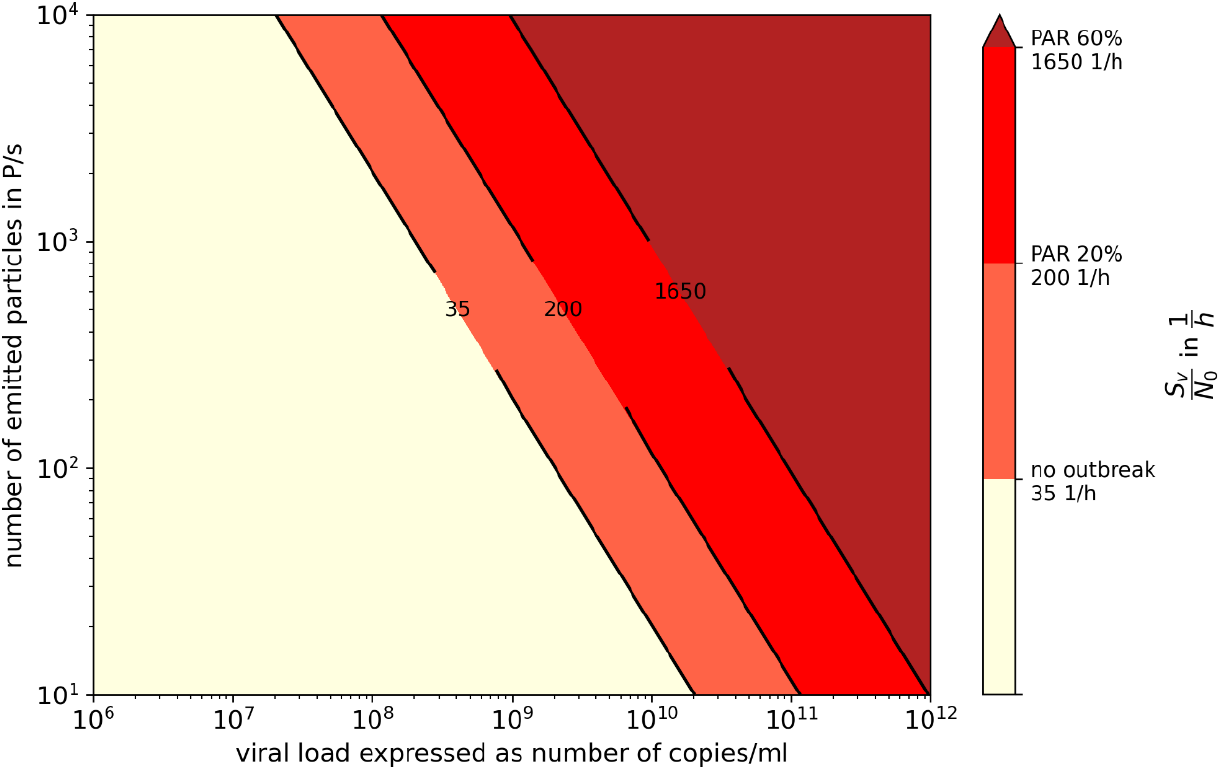
viral emission for different viral loads and particle emission rates with N_0_=67 viral copies, the attack rates found in the investigated outbreaks are shown with the different colors

**Figure 9:**
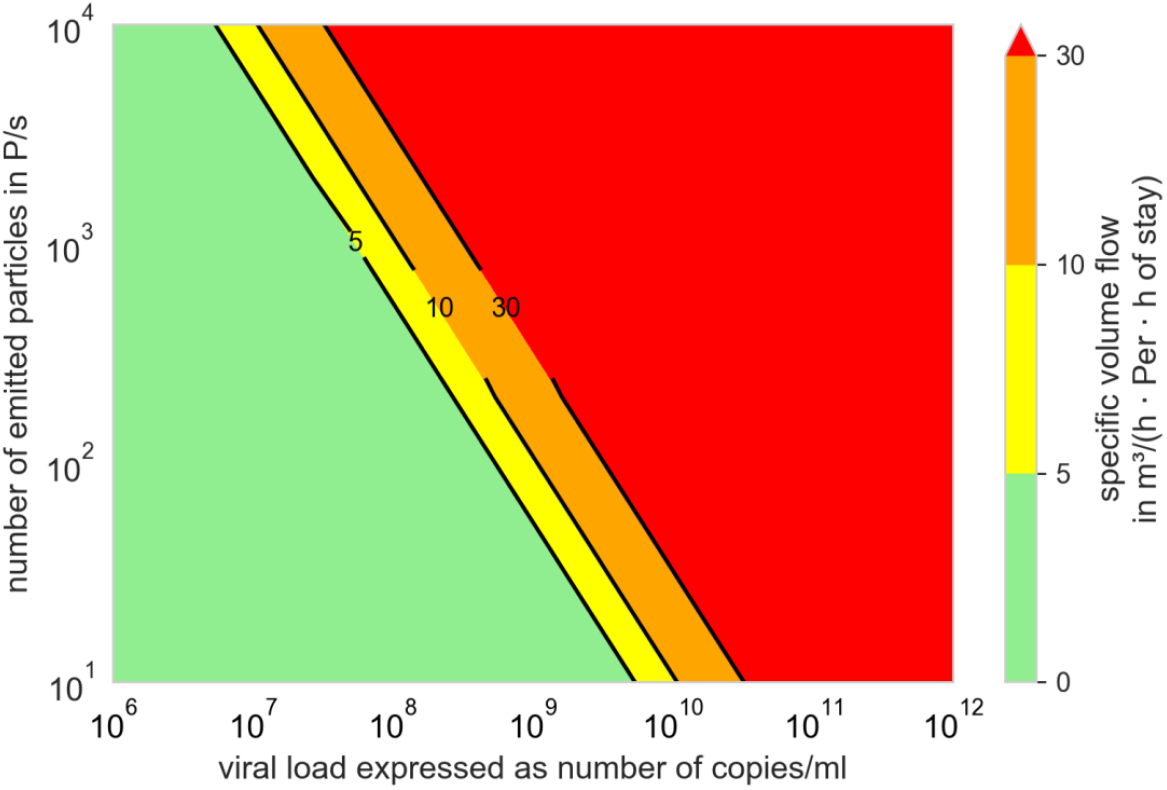
specific volume flow depending on the number of emitted, particles and the viral load to limit the number of newly infected persons to one, N_0_=67 viral copies, f_M_=1, Q_b,in_=Q_b,e_=0.54m^3^/h

### Comparison of prevention measures Ag testing, wearing masks, and increasing ventilation rate

As can be seen from equation (35), the VF (S_v_/N_0_) is dominant in PAR_S_. S_v_/N_0_ varies by a factor of 1,000 between a viral load of 1E+08 and 1E+11. The preventive measures (increasing the virus-free supply air volume, wearing masks, reducing the time of stay and their combination) in the specific situation have to be in a similar order of magnitude to actually prevent an outbreak. The comparison is performed with the simplified model, which is valid for lower PAR_s_ and in case of high PAR_s_ further measures should be implemented anyway, so that the actual value for higher PAR_s_ is not relevant. If the lower limit of the supplied virus free air volume flow is calculated to reach a CO_2_-concentration of 4000 ppm (common for not regularly performed window ventilation and longer stays) and the volume flow is increased until a lowered CO_2_-concentration of 1000 ppm is reached (complies with the normative recommendation for indoor air quality), the factor of change is 7 related to the air volume flow and the preventive impact. To wear a face mask will on average reduce the inhaled dose by 50 %, whereas a factor of 2 can be applied as well as for halving the time of stay in the room together with the infectious person. For a FFP2-mask the dose will, in average, be reduced by 80 %, whereas a factor of 5 can be applied.

From Figure 10 it can be seen, that even with a combination of different measures an outbreak cannot be avoided completely, and infections may occur, if the viral load is high enough.

**Figure 10:**
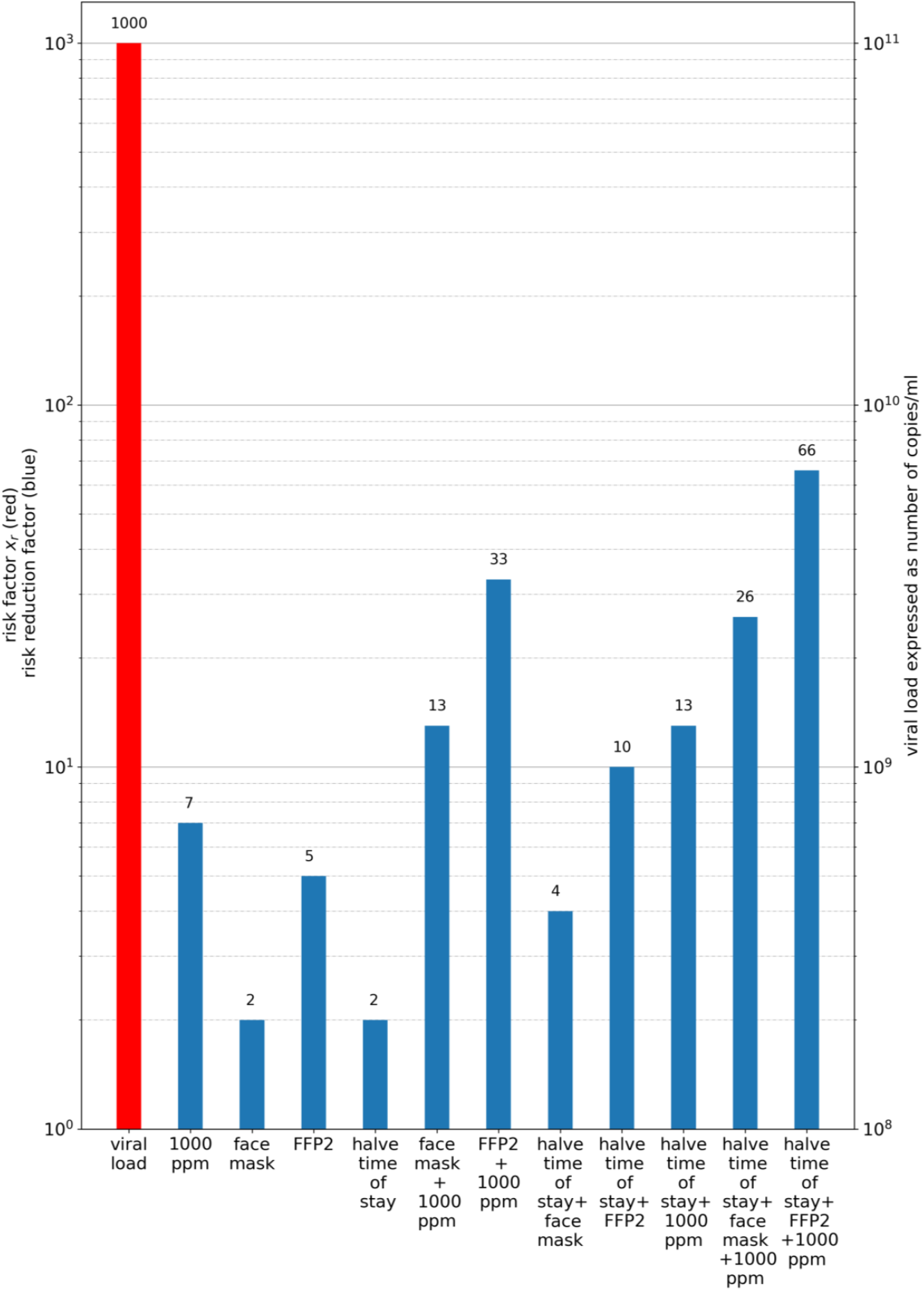
influence of different preventive measures on the risk of an outbreak. The red bar represents the viral load and the resulting risk factor (eq. 37). The blue bars illustrate different combinations of preventive measures in the form of a risk reduction factor (also according to eq. 37).

Only a small change in viral load with a factor of 10, e.g. from 1E+08 to 1E+09, could probably be compensated by wearing masks and ventilate regularly (Figure 10, blue bar: face mask + 1000 ppm). If it can be avoided, that the virus source enters the room, it is obvious, that this is the most effective preventive measure. Ag-tests can be of practical use, even if their sensitivity is limited for low and medium viral load.

## Discussion and Limitations

A dose-response-model to evaluate the risk of infection with SARS-CoV-2 was used to analyze twenty-five outbreaks in different situations (e.g. school, choir, meetings).

Although the viral load in the investigated outbreaks were unknown, our results strongly suggest that relevant transmission will take place when viral loads are high. Data of particle emission rate during various activities were used to infer virus emission rate, but this varies significantly among individuals (Figure 2). Even if the particle emission is significantly under- or overestimated, a viral load of at least 1E+08 viral copies/ml would have to be existent (see Figure 3 and Figure 4). Using this particle emission rate and size distribution, a number of aerosolized viruses is obtained as a function of viral load. It is assumed that the viruses at the site of aerosol generation in the body corresponds to the viral load in the swab, which remains to be proven.

The highest calculated value is found for a choir rehearsal with 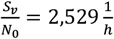 and is therefore within the range found by Buonanno et al [31]. The lowest value of 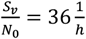 is found for a bus travel, where the activity and/or the viral load might have been low, which is comparable to the results of Buonanno et al [31] as well. The high emission during the choir rehearsal seems valid, because of the high particle emission rates while singing and the low air change rate. Furthermore, it has to be taken into consideration that the particle emission for different persons might be different for the same activity so the 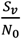 can be quite different, but still the main conclusion will remain.

Regardless of the claim for correct quantity of particle emission strength and viral load, it was shown that the specific person-related airflow rate is a practical quantity for evaluating ventilation-related measures, simply mathematically derived, see (24) - (32).

It has also been clearly demonstrated by us that preventive ventilation measures can prevent outbreaks due to aerosol transmission only in a narrow band. At high virus loads, the outbreak size can be reduced, but not prevented. Additional preventive measures are often necessary (Figure 5, Figure 9 and Figure 10).

If the viral load must be so high to cause infection due to aerosol transmission, then even using Ag Tests can be very effective measure to prevent infectious individuals from entering the room and causing an outbreak. Nevertheless, their efficiency depends on quite a lot of influencing factors, like the quality (sensitivity) of the test and the correct execution of the test. Even if studies showed that in general, most pupils are able to perform the Ag-tests correctly if instructed, a repetition on regular basis may e.g., result in less attention paid to correct testing. Also, if antigen tests are not carried out on a daily basis, the possibility exists that a person with a positive test result already exhibited transmissible virus loads the previous day.

However, different limitations as well regarding the model as the practical application have to be considered. First of all, different influencing factors (e.g. critical dose, decay rate of sedimentation as well as inactivation, size distribution and number of emitted particles) were assumed based on current knowledge, but whereas the transmission of SARS-CoV-2 is still ongoing, further knowledge may be gained from further research. The decay rates of sedimentation as well as inactivation can be influenced by the particle size distribution as well as the air temperature and humidity in the room. Furthermore, the analysis and model base on some general assumptions like ideal mixing of all particles within the room, an initial concentration of 0 virus copies/m^3^ and a supply of virus free air. An ideal mixing of all particles in the room also implies, that no separation into near and far field can be performed, whereas the concentration of virus laden particles near the person is probably higher than in the rest of the room. Also, the local concentration will differ regularly from the average concentration so that the local air quality index should be considered for investigations in detail [27, 28]. As a result, even at lower viral emission rate S_v_, infection can occur via aerosol, predominantly in the near field. As a third aspect the influence of VOCs is difficult to define. A higher transmission rate of new VOCs may result from different aspects, such as a change in critical dose, a change in viral load or a change in other measures. It also has to be kept in mind, that the investigated outbreaks documented with ARs between 4 % to 100 % have had mostly high ARs and therefore result in a high number of newly infected persons. Many transmission events have much lower ARs, so the results may over- or understate the true risk.

It could be shown that for viral loads smaller than 1E+08 viral copies/ml, aerosol transmission becomes unlikely if the distance is maintained. But it has to be considered that in some of the investigated cases, the range between the 25 % and the 75 %-percentile is quite high, which is because of insecure boundary conditions.

## Conclusion

1. **For an outbreak due to aerosol transmission to happen, high viral loads are required, which regularly occurs with the Delta variant**.
2. **Preventive measures such as wearing masks and rising ventilation cannot prevent an outbreak when virus loads are very high, but are useful to mitigate it**.
3. **The person-related air flow rate per hour of stay is the favorable indicator for evaluating the preventive effect of ventilation measures. According to our observation even volume flow rate and person related volume flow rate have more informative quality than the air change rate**.
4. **Instead of a CO**_**2**_ **concentration, the CO**_**2**_ **dose (integration of the difference to the outdoor air concentration) is suitable for defining a limit value that should not be exceeded**.
5. **With a simplified approach it is easy to compare different indoor situations and preventive measures regarding aerosol transmission**.
6. **Ag tests bring an effective additional value: they have a high sensitivity (detection rate) at virus loads of more than 1E+06 viral copies/ml and are therefore able to detect infectious persons with a chance to isolate them before entering a room for a longer stay**.

From the investigation of the outbreaks, it can be concluded that in all cases a viral load of at least 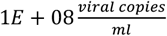 have been necessary to reach the observed attack rates. It can be seen that the viral emission of the infected person is the dominant influencing factor, but three further aspects (situational aspects, personal aspects of the susceptible persons and preventive measures) have to be considered to transfer a transmission of SARS-CoV-2 into a superspreading event. It can be concluded that solely a high activity (like singing or physical activity) does not necessarily result in high ARs (Choir Rehearsal Berlin 2) and that lower activity may also result in high ARs (e.g. School Hamburg 1) if other unfavorable conditions (e.g. high viral load, ineffective preventive measures) occur.

Nevertheless, a higher activity will result in a higher emission rate of particles and therefore in a higher concentration of virus laden particles in the room air. For transmission, aerosol production and viral load must always be considered together. A person considered infectious does not mean that aerosol transmission will also occur.

The comparison of different preventive measures demonstrates that these measures alone as well as in combination are able to reduce the rate of transmission, but for high viral loads still high infection rates will occur. Especially with the VOC Delta, for which an up to 1,000-times higher viral load was found for persons, there is a high risk of outbreaks and superspreading events even when the AHA+L rules are observed (German Abbreviation for masks, hand hygiene, social distancing and ventilation).

We showed, that to recommend an air change rate *λ*_*ACH*_ alone is not sufficient for infection prevention, whereas the person related volume flow of virus free supply air is a much more relevant parameter. The specific volume flow per person can be correlated with a CO_2_-concentration in the room, but whereas the number of inhaled virus laden particles increases with time, the CO_2_-concentration will reach a steady state concentration after a certain time, and does not change much anymore until the persons leave the room. In comparison the number of inhaled virus laden particles increases over time even if their concentration in the room stays constant. Therefore, to use a fix CO_2_-concentration as an indicator for the risk of infection has important limitations. Instead the CO_2_-dose (*ppm* ▪ *h*) can be used meaningfully and is easy to integrate in an infection risk monitoring system.

In case of a high number of susceptible persons (*N*_*pers*_ > 5) and low predicted attack rates (*PAR*_*s*_ < 20%) a simplified model was set up, which can be used to predict the influence of different measures on the risk of infection, to calculate the maximum number of persons or to calculate the necessary volume flow per person to avoid an infection of more than one person and is applicable to compare different indoor situations. Therefore, for high ARs the simplified model is not applicable, it is suitable that the model is used before outbreaks happen, where low ARs shall be intended. For retrospective analysis a more detailed model should be used.

High viral loads, which may result in a transmission via aerosol particles, can be found with antigen rapid tests. In this case the exposition would not occur (if the person does not enter the room at all) or is of short duration (if it the test is performed in the room as well).

Due to the fact, that the concentration of viral copies in the surrounding of an infected person is always higher and that there are insecurities regarding the tests as well as the transmission from or to vaccinated or recovered persons, a multilayer approach of preventative measures like wearing masks and increasing the air flow rate are necessary to lower the infection rate. For future outbreaks it would be helpful if all boundary conditions (e.g. volume flow, room size, time of exposure, etc.) and viral load at the moment of transmission are determined retrospectively. A summary of the necessary information for the boundary conditions is presented in the appendix.

## Data Availability

All data produced in the present work are contained in the manuscript.

## Declarations

The authors received no specific funding for this work.

The authors declare no competing interests.

The authors declare that they followed the appropriate research guidelines.

## Appendix

### Calculation of the conversion factor from number to volume of the particles

In different studies, the particle emission rate for different activities was measured. The results found by some of the authors of this paper can be seen in Table 2. It has to be mentioned that the particles were measured about 0.81 m behind the mouth of the subjects. Therefore, it was assumed that they already reached equilibrium diameter, when entering the particle counter. To take this into consideration the evaporation has to be considered. Regarding [40] it is considered that the equilibrium diameter is between 33 and 50% of the original diameter.

To calculate the conversion factor from the particle emission rate to the volume of the particles first of all the volume of a particle has to be calculated. Spherical particles were assumed for this calculation. Furthermore, for each size class the minimum diameter is assumed to be most representative, because more smaller than larger particles were found. With this assumption the volume of the particles in the different size classes can be calculated with a shrinking factor of 2 so a particle with a diameter of 0.3 μm, was calculated with an initial diameter of 0.6 μm.

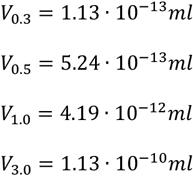

For adults, the conversion factor is calculated with the following distribution of size classes when speaking: 57% 0.3-0.5 μm, 23% 0.5-1.0 μm, 18% 1.0-3.0 μm and 2% larger than 3.0 μm.

The average emission of volume by exhaling one particle can therefore be calculated using equation (38) where i denotes the size class, f_i_ the proportion of particles in the size class and V_i_ the volume of particles in this size class.

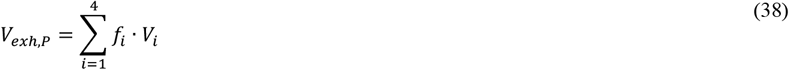

For breathing the exhaled volume of particles is therefore 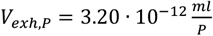. The conversion factor shall further include the conversion from seconds to hours and is therefore 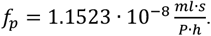.

### Necessary Boundary Conditions to retrospectively investigate outbreaks

For the investigation of outbreaks, different boundary conditions are necessary. The highlighted are elementary, the others optional. They are summarized in the following:

1. virus related aspects
  a. How many people got infected by the virus?
  b. How many people attended the event?
  c. How many people were vaccinated or recovered from an infection?
  d. Has it been defined which type of virus caused the infection? If yes, which one?
  e. How high was the virus load during the infection event?
2. room-related aspects
  a. How big is the room in which the event took place (area, volume)?
  b. Was the room ventilated mechanically?
    i. Which volume flow or air change rate was available?
  c. Was the room ventilated by window opening?
    i. How often and for how long have the windows been opened?
    ii. Is there anything known, about the outdoor conditions on that day? (temperature, wind speed, wind direction)
3. event-related aspects
  a. How long took the event?
  b. Did all attendees stayed in the room together for the whole event? Otherwise specification, which part left for how long
  c. What was the main activity of the infectious person?
  d. What was the main activity of the susceptible persons?
  e. Have there been any additional preventive measures (e.g. face masks,…)?
  f. Were the persons at fixed positions during the stay? What were the approximate positions of the persons?

### Boundary Conditions of the investigated Outbreaks

For twenty-five different outbreaks information was available or have been assumed.

Four different choir rehearsals are considered. All choirs rehearsed in unventilated or just slightly ventilated rooms. The number of attending persons ranged from 25 to 77 persons and the room size from 135 to 1720 m^3^. The duration of the rehearsals differed just slightly between 2 and 2.5 h.

In the Choir Rehearsal Berlin 2 the previously infected person was the choir director and was therefore not singing the whole time. Therefore, a mixture of singing and speaking was assumed [39].

In the French Choir three members of the choir got tested positive for SARS-CoV-2 within the next days after the rehearsal [73]. Although one member had symptom onset the day after the rehearsal it seems reasonable to assume that only one person emitted the virus laden particles.

The second group consists of outbreaks either involving loud speaking or intense physical activity like sports or heavy working. For three out of the four outbreaks little information regarding the ventilation system in the room are available, so that typical values have been assumed, either by the authors of this study (Hawaiian fitness class [74]) or by authors of earlier studies (Korean Call Center and Korean fitness center [30]). Solely for the German Slaughterhouse measurements of the volume flow were performed afterwards [75].

For the working places (call center [76] and slaughterhouse [75]) the time of stay was a working day (8 hours) whereas for the fitness class the duration of stay was significantly smaller (approximately 1h or less [74, 77]).

Two outbreaks with normal office activity, but relatively high attack rates are considered as well [78, 79]. One outbreak happened during a school day (4.5 h) in a ventilated classroom [78] and the other during a court session (3 h) in an unventilated courtroom [79]. Both rooms had a similar size. Furthermore, an outbreak in a restaurant was included into this category. It is one of the first documented outbreaks related to airborne transmission of SARS-CoV-2. The boundary conditions, mainly room volume and ventilation were assumed by Li et al [1].

In addition, some outbreaks related to public transport were found [80, 81, 82, 83]. One outbreak happened in an aircraft during a 11 h flight with 217 passengers and the other in buses of different sizes (3 to 68 passengers and 16 to 71m^3^). The bus travel lasted between 1 and 2.5 h.

Finally, two meetings as well as six outbreaks in school classes, which were investigated and provided by local health authorities, or the Robert-Koch-Institute are considered. For the outbreaks, the air change rate has either been assumed afterwards depending on the ventilation habits and the temperature and wind speed on the day of outbreak or known from existing ventilation systems. The room volume, the number of attending persons as well as the time of stay was investigated by the health authorities together with the attack rate reported for this outbreak.

The index person in two of the school cases (School Hamburg 3 and School Hamburg 4) wore a mask (f_M_=0.7) and noted some symptoms, whereas he reduced speaking intensity.

### Comparison of different situations

To compare different situations with the simplified approach a viral load of 1E+08 viral copies/ml was used. The situation in the supermarket with mask, like it is currently common, where *R*_*s*_ = 1 is used as the reference. In all situations the duration of stay is longer than in the supermarket, but in the theater/cinema scenario the risk is lower than in the supermarket. In all other situations the risk is higher and if the duration of stay is much longer (office or school) the risk is significantly higher and further measures (reduction of the number of persons in the room, wearing masks) are necessary to reduce the risk.

